# Blood and tissue correlates of steroid non-response in checkpoint inhibition-induced immune-related adverse events

**DOI:** 10.1101/2024.12.04.24318205

**Authors:** Mick J.M. van Eijs, M. Marlot van der Wal, Hedi-Britt Klotškova, Noël M.M. Dautzenberg, Mark Schuiveling, Rik J. Verheijden, Fiona D.M. van Schaik, Bas Oldenburg, Stefan Nierkens, the UNICIT Consortium, Karijn P.M. Suijkerbuijk, Femke van Wijk

## Abstract

High-dose steroids constitute the cornerstone of first-line treatment for immune-related adverse events (irAEs) associated with immune checkpoint inhibitors, but compromise antitumor immunity. A deeper understanding of irAEs and their response to steroids can contribute to more targeted irAE management regimens. We took a multi-omics approach to identify blood- and tissue-based predictors of steroid response and to explore underlying mechanisms of steroid non-response in irAEs. In the blood, steroid non-response correlated with trends for elevated Tc1/Tc17 CD8^+^ T cells and serum interleukin (IL)-17, IL-6, IL-12 and IL-23 prior to initiation of steroids, along with persistent (CD8^+^) T cell proliferation and activation after start of steroids. A remarkably fast decrease in inflammatory gene signatures and lymphocyte infiltration was observed in colitis tissue of steroid responders obtained within 24h after initiation of steroids. Peripheral T cell PD-1 receptor occupancy was not associated with steroid response. Colitis tissue of steroid non-responders was enriched for activated CD4^+^ memory T cells and a pronounced type 1/17 immune response. Together, our findings suggest rapid immunological effects of steroids in circulating cells and irAE-affected tissue and support that an enhanced type 1/type 17 response is associated with steroid non-response in irAEs.

## Introduction

Immune checkpoint inhibition (ICI) has revolutionized the field of oncology, with expanding indications across tumor types and in both early and advanced disease stages.^1^ Unfortunately, ICI comes with severe immune-related adverse events (irAEs) in up to 60% of patients treated with combined anti-programmed death-1 (anti-PD-1) plus anti-cytotoxic T-lymphocyte-associated protein-4 (anti-CTLA-4).^1^ Most severe irAEs do not resolve spontaneously without immunosuppressive treatment and can become chronic, or even fatal, if left untreated.^1^ Clinical guidelines empirically advise to treat irAEs, depending on Common Terminology Criteria for Adverse Events (CTCAE) severity grade, with moderate-to-high doses of systemic steroids in first line (typically 1 or 2 mg/kg daily prednisone-equivalence for some grade 2 and most grade 3-4 irAEs).^2-4^ If symptoms do not improve within 3-5 days, escalation to higher-dosed steroids and/or initiation of second-line immunosuppression are recommended.^2-4^

Data from our group illustrate that the current empirical approach is not optimal, as we showed that both high peak-dose steroids and second-line immunosuppression are independently associated with worse overall and cancer-specific survival in patients with melanoma treated for irAEs.^5-8^ These findings warrant investigation of alternative strategies to limit cumulative exposure to immunosuppressants by timely introducing tailored second-line agents or skipping steroids dose escalation treatments. On the other hand, we currently lack mechanistic cues and clinical predictors for steroid unresponsive types of inflammation, which is urgently required to design personalized alternative treatment regimens.^1^

Here, we applied a multi-omics approach to find blood- and tissue-based predictors for response to steroids, longitudinally evaluated effects of steroids in peripheral blood immune responses and explored potential mechanisms driving steroid non-response in irAEs.

## Results

In March 2024, a total of 535 patients had been prospectively enrolled in the UNICIT biobank, which includes patients with solid tumors undergoing ICI treatment at University Medical Center Utrecht, the Netherlands.^9^ During a median follow-up of 23.6 months (95% confidence interval [CI]: 20.4-26.2), 156 unique patients developed 197 grade ≥2 irAEs requiring ≥0.5 mg/kg/day corticosteroids. Of those 156 patients, blood was collected at the onset of irAEs before initiation of immunosuppressive treatment in 87 patients (55.8%). Response to steroids and subsequent treatment lines for irAEs was recorded for all patients. Steroid non-responders can be categorized as steroid-resistant, steroid-refractory or steroid-dependent, depending on their initial response to steroids and relapse of symptoms upon steroid tapering.^10^ In this study, we considered patients “steroid non-responders” when they required any second-line immunosuppression to achieve complete remission of irAEs before steroids had been fully tapered. A complete overview of the study design, including number and type of samples used and analyses performed, is in **Figure 1**.

**Figure 1.**
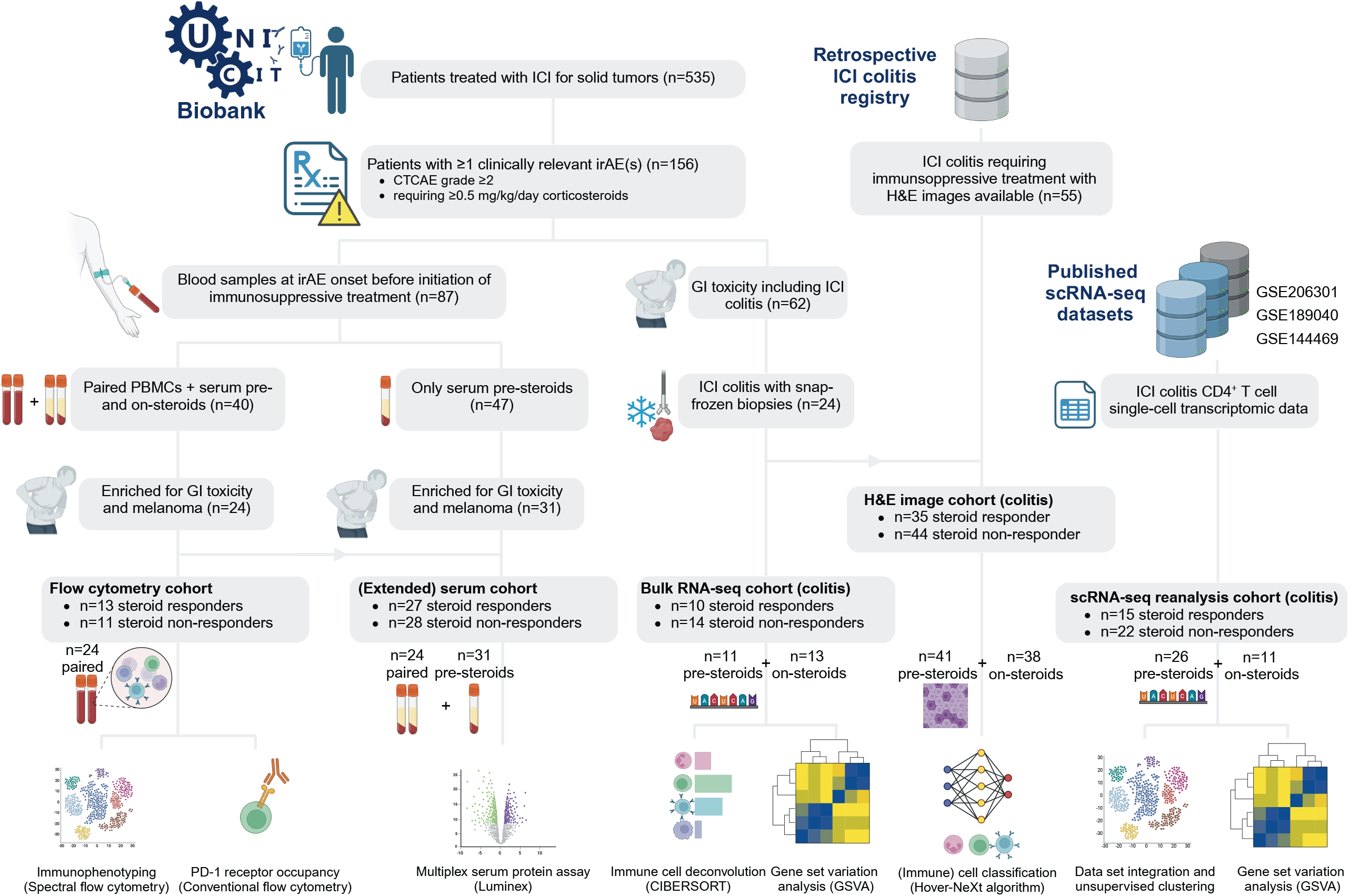
Study design. Overview of patients included in this study, type and number of samples used and the sources for all samples and data. For all patients (samples), the response to steroids and timing of sampling (pre-steroids, on-steroids or paired pre- and on-steroids) is shown. Abbreviations: CTCAE denotes ‘Common Terminology Criteria for Adverse Events’, GI ‘gastro-intestinal’, H&E ‘hematoxylin and eosin’, ICI ‘immune checkpoint inhibitor’, irAE ‘immune-related adverse event’. Detailed patient characteristics are in **Supplementary Tables 1-4**.

From all UNICIT patients who received ≥0.5 mg/kg/day corticosteroids for irAEs, we first selected patients with paired peripheral blood mononuclear cells (PBMC) and serum samples prior to and following initiation of steroids for irAEs (**Supplementary Table 1**). We assessed abundance and functional phenotypes of immune cell subsets with spectral flow cytometry in pre- and on-steroids samples together. Annotation of 50 unsupervised clusters yielded 15 peripheral blood immune cell subsets (**Figure 2a, Supplementary Figure 1a**).

**Figure 2.**
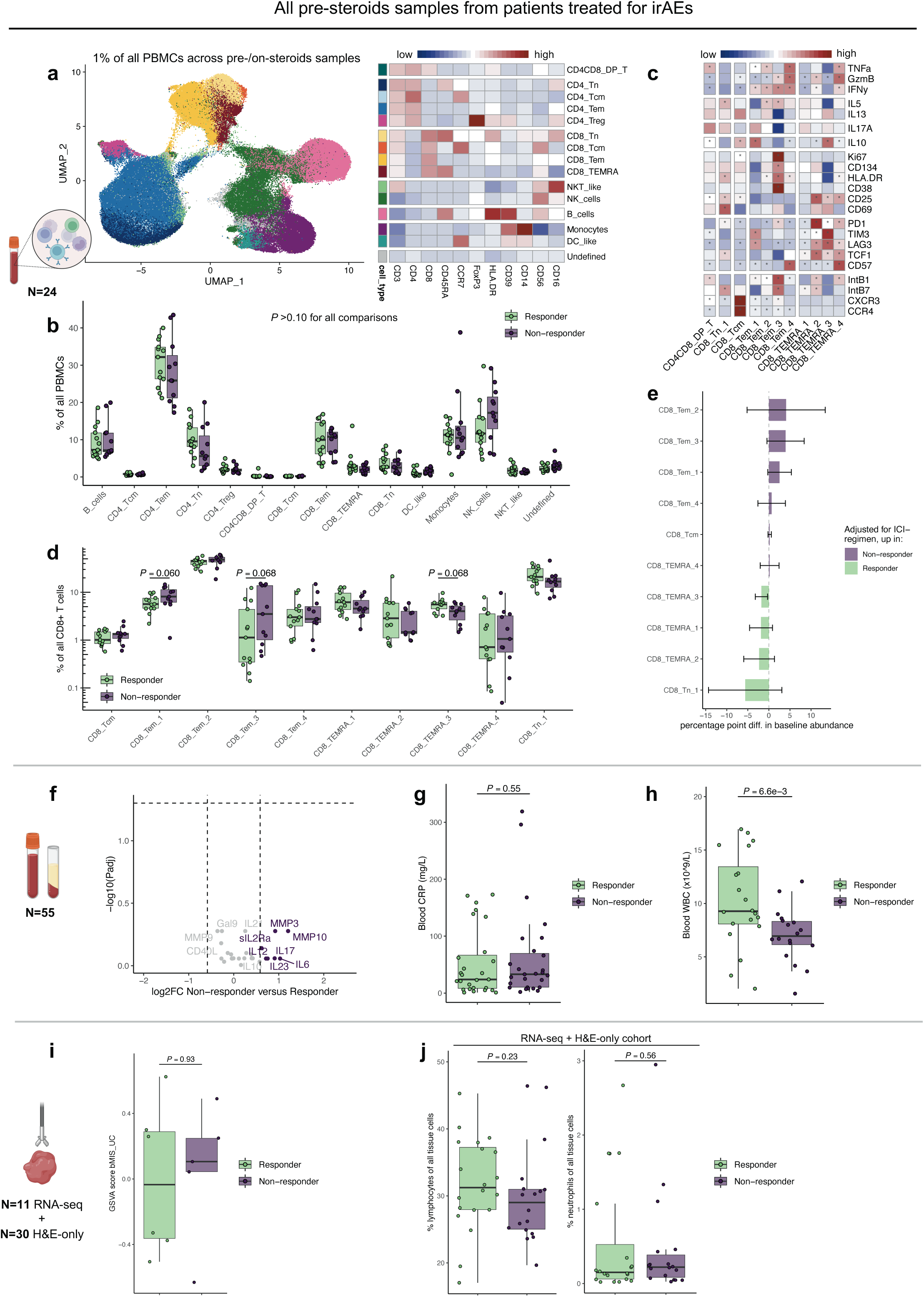
Blood- and tissue-based prediction of steroid response. Unadjusted P values are displayed, unless otherwise specified. **a** Clustering of PBMCs based on UMAP embeddings obtained in a 1% random subset of the entire dataset (including all time points), overlaid with annotated FlowSOM-based clusters, along with the expression heatmap displaying median expression of lineage-defining markers in FlowSOM-based clusters. For the lack of B-cell-specific markers, B cells were annotated by exclusion and based on HLA-DR, CD39 and CD45RA positivity. **b** Boxplots displaying pre-steroids abundance of PBMC subsets by response to steroids. **c** Expression heatmap displaying median expression of functional markers in CD8^+^ T cell subsets, grouped from top to bottom as Th1-related, Th2-related, Th17-related, Immunosuppressive, Proliferation/activation, Dysfunction/exhaustion and Homing/trafficking. Except CD57 and HLA-DR, these markers were not used as clustering parameters in FlowSOM. * Significant marker enrichment at P_adj_<0.05. **d** Boxplots displaying pre-steroids abundance of CD8^+^ T cell subsets by response to steroids. **e** Relative pre-steroids abundance of CD8^+^ T cell subsets by response to steroids, adjusted for ICI treatment type. Linear regression coefficients are shown with whiskers indicating 95% confidence intervals. **f** Volcano plot displaying difference in pre-treatment concentration of 26 serum proteins measured by multiplex assay in responders versus non-responders in the full extended cohort. **g**,**h** Boxplots displaying pre-treatment (**g**) C-reactive protein (CRP) and (**h**) blood leukocytes (WBC), by response to steroids in the full extended cohort. CRP and WBC were missing in 1 and 17 patients, respectively. **i**,**j** Boxplots displaying pre-steroids (**i**) molecular inflammation assessed by gene set variation analysis using the bMIS_UC score and (**j**) percentages lymphocytes and neutrophils of all tissue cells, identified by HoVer-NeXt, in H&E slides of ICI colitis, by response to steroids.

### Blood- and tissue-based prediction of steroid response

We first aimed to identify predictors for steroid response in blood by analyzing pre-steroids samples only, collected median 1 day (IQR: 0-3 days) before initiation of steroids. At irAE onset, before steroids had been initiated, none of the 15 immune cell subsets predicted steroid response (**Figure 2b**), irrespective of ICI type (**Supplementary Figure 1b,c**). We then focused on different T cell subsets, by assessing expression of functional markers in original unsupervised clusters (**Figure 2c** for CD8^+^ T cells, **Supplementary Figure 1d** for CD4^+^ T cells). None of the detailed T cell clusters was predictive of steroid non-response after multiple testing correction. However, based on unadjusted *P* values, a trend for relatively higher CD8^+^ Tem cluster 1 (Tc17-like; IL17A^hi^IFN-γ^hi^ but granzyme B^lo^) and CD8^+^ Tem cluster 3 (activated, proliferative, cytotoxic Tc1-like, equipped for gut homing; integrins β1/β7^hi^,^11^ granzyme B^hi^IFN-γ^hi^Ki67^hi^CD38^hi^HLA-DR^hi^ but IL17A^lo^), and lower exhausted CD8^+^ T_EMRA_ cluster 3 (PD-1^hi^LAG-3^hi^TIM-3^hi^ but granzyme B^lo^IFN-γ^lo^TNF-α^lo^) in steroid non-responders was found (**Figure 2d**). These trends remained after adjustment for ICI type (**Figure 2e**). Pre-steroids CD4^+^ T cell clusters were not predictive of steroid response, but we could confirm our previous finding that ICI type was associated with the extent of CD4^+^ effector memory proliferation (**Supplementary Figure 1e**,**f**).^9^

Subsequently, we measured serum concentration of 26 analytes in pre-steroids samples of the same patients, extended with pre-steroids samples obtained ≤4 days prior to steroids initiation from 31 extra patients (**Supplementary Table 2**). In addition, complete blood counts with differential and C-reactive protein (CRP) concentrations were available from regular diagnostics in most patients. None of the analytes was differentially abundant pre-steroids in steroid non-responders compared to responders at *P*_adj_<0.05, but non-responders showed a trend of elevated (>1.5×) serum interleukin (IL)-6, IL-17, IL-12, IL-23, matrix metalloproteinase-3 (MMP-3), MMP-10 and soluble IL-2-receptor (**Figure 2f**). MMP-3 derives from various connective and visceral tissues and is strongly inducible by IL-17,^12-14^ suggesting type 17 skewing in tissue of non-responders. Although IL-6 was numerically twofold higher in steroid non-responders than responders, pre-steroids CRP concentration, induced among others by IL-6,^15^ was equal between both groups (**Figure 2g**). Steroid responders featured higher total leukocyte count (**Figure 2h**), including elevated eosinophils, neutrophils and lymphocytes before initiation of steroids (**Supplementary Figure 1g-i**). Relative leukocytosis may reflect the acute phase of irAEs, which could indicate that irAEs respond better to steroids early in the disease course.

Finally, we selected all 24 patients from the biobank with histologically confirmed ICI colitis and snap-frozen biopsies obtained either before or after initiation of steroids (**Supplementary Table 3**). Bulk RNA isolated from whole colon biopsies was sequenced and absolute abundance of 22 immune cell types was deconvoluted with CIBERSORT.^16^ First, we only analyzed samples from the 11 patients that were steroid-naïve at endoscopy and had sufficiently reliable deconvolution results (at *P*_permutation_<0.05). Before initiation of steroids, non-responders had more activated memory CD4^+^ T cells in ICI-type-adjusted analysis (**Supplementary Figure 1j**). Pre-steroids extent of mucosal inflammation was assessed molecularly by gene set variation analysis (GSVA) using a biopsy molecular inflammation score developed in ulcerative colitis (bMIS_UC).^17^ Higher bMIS_UC indicates more inflammation, but the bMIS_UC was not different between steroid responders and non-responders (**Figure 2i**).

To accommodate the limited sample size of snap-frozen tissue before steroids initiation, we extended this cohort with 55 ICI colitis patients (including 30 steroid-naïve patients) from whom digital images of hematoxylin and eosin (H&E) stained colon slides were available (**Supplementary Table 4**). Nuclei were segmented and connective tissue, epithelial cells, lymphocytes, plasma cells, neutrophils and eosinophils were annotated with the HoVer-NeXt algorithm specifically trained on colon tissue data.^18^ When analyzing steroid-naïve samples only, no differences were observed in total lymphocyte or neutrophil infiltration in relation to steroid response (**Figure 2j**).

In conclusion, steroid non-response correlated with trends for increased circulating percentages of Tc1- and Tc17-like CD8^+^ T cells and elevated serum IL-12, IL-23, IL-6 and IL-17 before initiation of steroids. In tissue, activated CD4^+^ T cells, but not total lymphocytes, neutrophils or molecular inflammation were associated with steroid non-response.

### Changes in blood and tissue after initiation of steroids in relation to response

To explore longitudinal differences in steroid responders versus non-responders, we analyzed the change in PBMC subsets and serum proteins in paired pre- and on-steroids samples. On-steroids samples were collected median 10 days (IQR: 6-17) after initiation of steroids, but before initiation of second-line immunosuppression in non-responders. Time between pre- and on-steroids samples was not significantly different between responders and non-responders (**Supplementary Table 1**). No changes of cell subsets or serum proteins after initiation of steroids were observed upon multiple-testing correction (**Figure 3a-e, Supplementary Figure 2a**). However, based on uncorrected *P* values we found persistently high CD8^+^ Tem cluster 3 (activated, proliferating Tc1-like) cells in steroid non-responders, whereas this cluster decreased median twofold in steroid responders (**Figure 3b,c**). Longitudinal serum analysis showed significant associations with steroid non-response based on unadjusted *P* values for persistently high or increasing levels of chemokines (C-X-C motif) ligand 10 (CXCL10), C-C motif (CC) ligand 4 (CCL4), CXCL13, A Proliferation-Inducing Ligand (APRIL) and MMP9 (**Figure 3d**). Particularly CXCL13, associated with germinal center activity, and APRIL, indicative of B cell activation, showed increases after initiation of steroids in non-responders (**Figure 3e**). Together, these data indicate persistent (CD8^+^) T cell activation and proliferation, germinal center activity and antibody responses in steroid non-responders.

**Figure 3.**
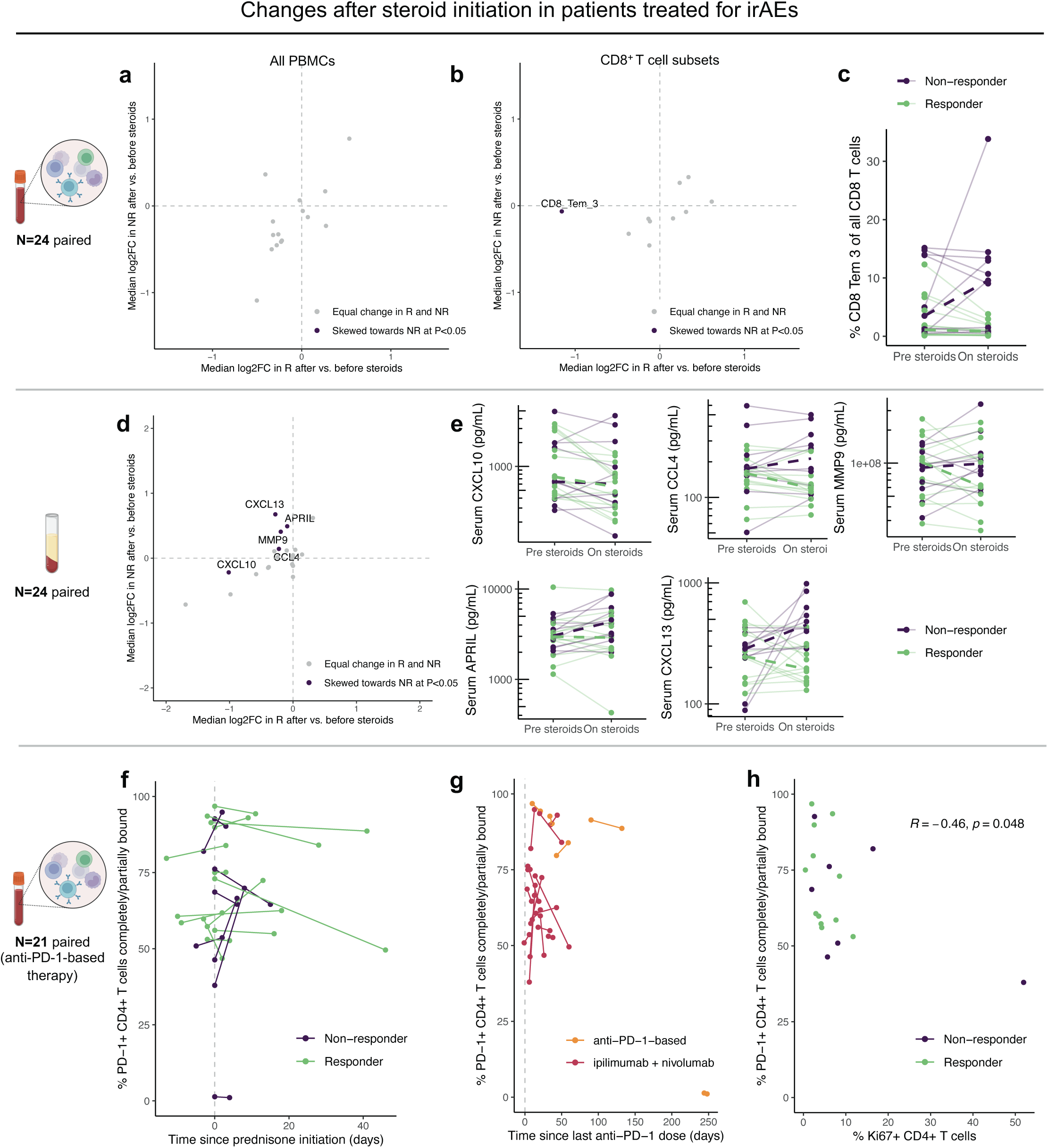
Changes in blood after initiation of steroids and PD-1 receptor occupancy in relation to steroid response. **a**,**b** Median log_2_ fold-change of (**a**) all PBMC and (**b**) CD8^+^ T cell subsets after initiation of steroids, in steroid responders (R; x-axis) versus steroid non-responders (NR; y-axis). Unadjusted P values <0.05 for the difference in fold-change between responders and non-responders are indicated. “Skewing towards” an index group can constitute a relative increase in that index group, a relative decrease in the comparator group, or a combination of both. **c** Spaghetti plot displaying the change in CD8^+^ Tem 3 cells for individual patients, colored by response to steroids. Dashed lines indicate median trends. **d** Same as (**a**,**b**), but displaying on-steroids change in serum protein concentration. **e** Same as (**c**), but displaying on-steroids change in CXCL10, CCL4, MMP9, APRIL and CXCL13. **f**,**g** Scatter plots displaying the percentage of PD-1^+^ CD4^+^ T cells completely or partially bound by anti-PD-1 therapeutic antibody in relation to (**f**) duration of steroids or (**g**) time since last anti-PD-1 administration, by (**f**) response to steroids or (**g**) ICI treatment type, for paired pre-/on-steroids samples. No formal statistical tests were performed. **h** Scatter plot displaying the percentage of PD-1^+^ CD4^+^ T cells completely or partially bound by anti-PD-1 therapeutic antibody in relation to CD4^+^ T cell proliferation for pre-steroids samples only. One patient with late-onset toxicity >200 days after the final ICI dose was excluded from this analysis.

Building on observations that anti-PD-1-bound T cells preferentially proliferate in the context of irAEs,^19,20^ we hypothesized that persistent activation and proliferation of peripheral blood T cells in non-responders would be indicated by a higher proportion of PD-1 receptor bound by therapeutic antibody. In contrast to our hypothesis, neither pre-steroids PD-1 receptor occupancy (RO), nor change in PD-1 RO after initiation of steroids was associated with steroid response (**Figure 3f, Supplementary Figure 2b**). As expected, PD-1 was completely unbound in a patient with late-onset toxicity >200 days after the last dose of ICI (**Figure 3g, Supplementary Figure 2c**). Surprisingly, PD-1 RO was lower in patients treated with combined ipilimumab plus nivolumab. Since combination ICI is associated with increased (CD4^+^) T cell proliferation, we suspected that rapid turnover of (bound) PD-1 receptor complexes in cycling cells could potentially distort measurements of PD-1 RO. This was supported by lower (apparent) PD-1 RO with higher CD4^+^ T cell proliferation (**Figure 3h, Supplementary Figure 2d**). However, internalization of drug-bound PD-1 did not increase over time in patients with lowest compared to highest PD-1 RO (**Supplementary Figure 2e,f**).

Next, we turned from blood to tissue and analyzed molecular inflammation and immune cell infiltration in colon biopsies from respectively 24 (with bulk RNA-seq data) and 79 (with H&E histological images) ICI patients that underwent endoscopy at different timepoints before or after initiation of steroids. Although no significant difference of bMIS_UC inflammation score was observed at baseline, lower bMIS_UC inflammation scores were observed one day after initiation of steroids in responders compared with non-responders, who exhibited high mucosal inflammation regardless duration of steroids (**Figure 4a**). This may reflect the clinical observation that some ICI colitis patients report symptom improvement within several hours to a day after initiation of steroids. Since pre-steroids molecular inflammation scores are unknown for responders who underwent endoscopy the day after initiation of steroids, confounding by (lower) severity should be considered. However, all three responders who underwent endoscopy at day 1 after steroids had grade 3 colitis, necessitating immediate steroidal treatment before endoscopic evaluation. In the extended cohort, comprising all 79 ICI colitis patients treated with steroids, a similar pattern was observed, with significantly lower lymphocyte infiltration (but not neutrophil infiltration) in steroid responders within the first 24h of treatment versus baseline (**Figure 4b**).

**Figure 4.**
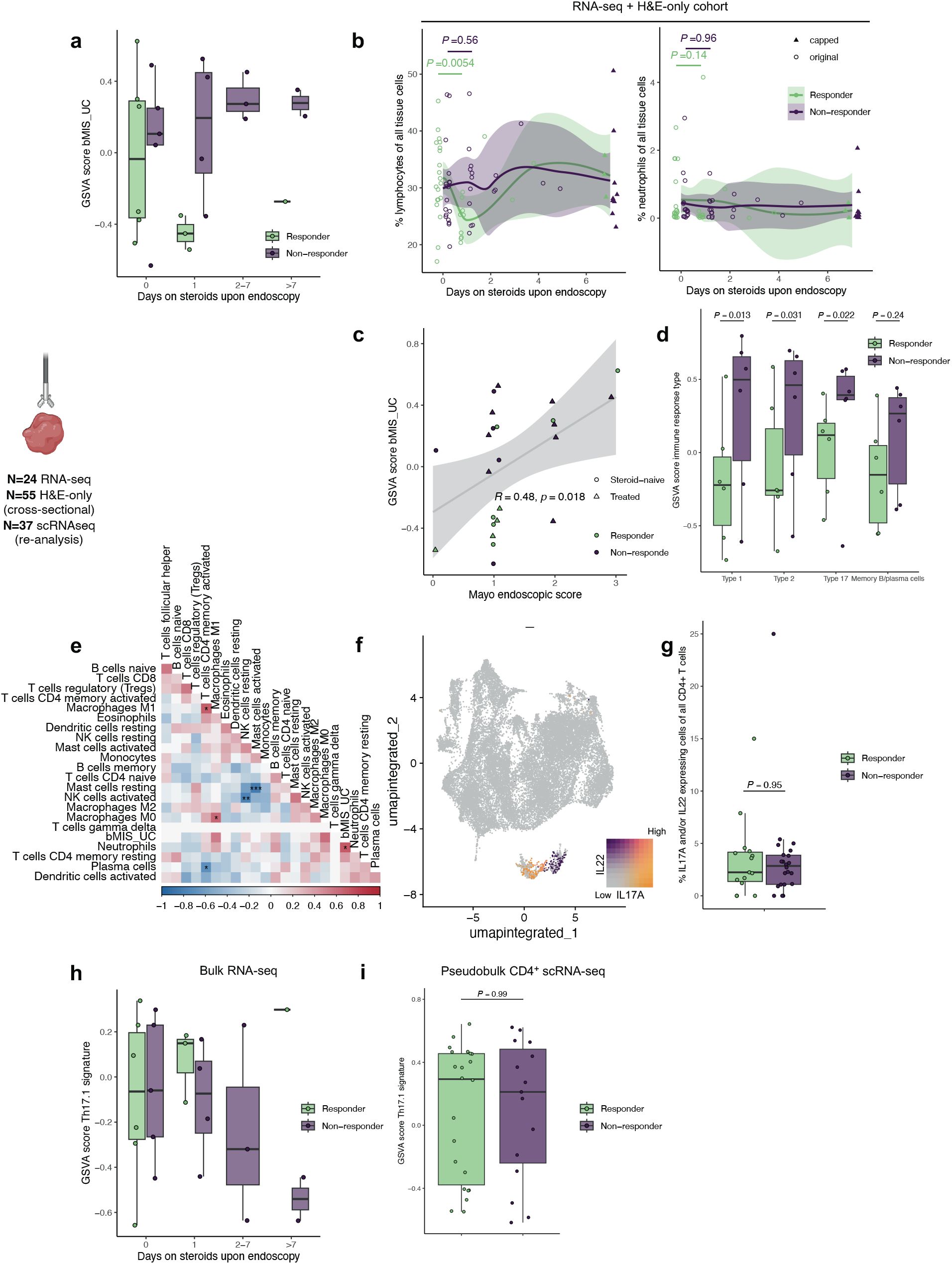
Tissue correlates of steroid non-response and exploration of underlying mechanisms. Unadjusted P values are displayed, unless otherwise specified. **a** Boxplots displaying molecular inflammation assessed by gene set variation analysis using the bMIS_UC score, including all biopsies of patients treated with steroids, stratified by duration of steroids upon endoscopy. No formal statistical tests were performed. **b** Scatter plots of percentages lymphocytes and neutrophils of all tissue cells in relation to duration of steroids upon endoscopy, using all H&E slides from ICI colitis patients treated with steroids. Trends in leukocyte infiltration in relation to duration of steroids are visualized by locally estimated scatterplot smoothing (LOESS). Datapoints with duration of steroids >7 days were winsorized at 7 days (indicated by solid triangles). Pre-versus 1-day on-steroids samples were compared by unpaired Wilcoxon tests, separately for steroid responders and non-responders. **c** Scatter plot displaying molecular inflammation (by GSVA bMIS_UC score) in relation to Mayo endoscopic score, with Pearson’s correlation with linear regression line and 95% CI. **d** Boxplots displaying GSVA scores for three immune response type-specific and one memory B/plasma cell gene sets in all biopsies (pre- and on-steroids samples combined), stratified by steroid response group. **e** Spearman’s correlation matrix of 22 deconvoluted immune cell types and bMIS_UC score in all samples with sufficiently reliable deconvoluted compositions (at P_perm_<0.05); *P_adj_<0.05, **P_adj_<0.01, ***P_adj_<0.001. **f** UMAP visualization displaying IL17A and IL22 expression in reanalyzed transcriptomic data of tissue CD4^+^ T cells in ICI colitis. **g** Boxplot displaying percentage IL17A and IL22 expressing cells of all CD4^+^ T cells in same patients as (**f**), by response to steroids. **h** Boxplots displaying Th17.1 signature enrichment in all bulk RNA-seq biopsies, stratified by requirement for immunosuppressive treatment and duration of steroids upon endoscopy. No formal statistical tests were performed. **i** Boxplots displaying Th17.1 signature enrichment in pseudobulk transcriptomic data of CD4^+^ T cells, by response to steroids.

In summary, steroid responders differ from non-responders in that peripheral blood (CD8^+^) T cell proliferation and activation is dampened, at least within several days of treatment, but possibly sooner. Persistent T cell proliferation in non-responders could not be linked to the extent of PD-1 receptor occupancy. Cross-sectional tissue data suggest that mucosal inflammation in ICI colitis may improve within 24h after treatment initiation in steroid responders.

### Exploration of mechanisms driving steroid non-response

Finally, we investigated immunological mechanisms in tissue underlying steroid response. Molecular inflammation correlated with the Mayo endoscopic score (MES), but provided a more granular impression of inflammation, particularly for endoscopically milder cases (**Figure 4c**). Since activated CD4^+^ T_mem_ cells were enriched in pre-steroids tissue of non-responders, we wondered if a specific T-helper response would underly steroid non-response. When using immunity-type-specific gene sets for GSVA in steroid-naïve and steroid-exposed biopsies combined, we found that type 1, type 2 and type 17 signatures were all significantly enriched in steroid non-responders, but type 1 and type 17 responses best differentiated between non-responders and responders (**Figure 4d**). Additionally, neutrophils were the only cell type that correlated significantly with “overall” mucosal inflammation quantified by the tissue molecular inflammation score (bMIS_UC) (**Figure 4e**). Therefore, we hypothesized that Th17 cells, which recruit neutrophils, could be drivers of steroid non-response and would be enriched in colitis tissue of steroid non-responders. To test this, we reanalyzed single-cell transcriptomic data from CD4^+^ T cells isolated from colon biopsies of 37 patients with ICI colitis from three studies.^20-22^ All patients who had been administered second-line immunosuppression were considered steroid non-responders. All studies comprised both steroid responders and non-responders; 70.3% of patients were steroid-naïve upon endoscopy. After correction of study-specific batch effect (**Supplementary Figure 3a,b**),^23^ unsupervised clustering yielded 25 CD4^+^ T cell clusters (**Supplementary Figure 3c**). Clusters representing Th17 and Th22 cells were identified based on expression of *IL17A* and *IL22* (**Figure 4f**). In contrast to our hypothesis, Th17/Th22 cells, as well as other CD4^+^ T cell clusters, were equally abundant in tissue of responders and non-responders (**Figure 4g, Supplementary Figure 3d**). This could be explained by subsets of Th17 cells differing in their pro-inflammatory capacity, with pathogenic Th17.1 cells particularly associated with steroid resistance in autoimmune disease.^24^ However, steroid responder and non-responder biopsies were also equally enriched for a published Th17.1 signature,^24^ both in bulk tissue and pseudobulk CD4^+^ T cell transcriptomic data (**Figure 4h,i**). In fact, CD4^+^ T cells from responders and non-responders were transcriptionally virtually identical, with no single gene differentially expressed at *P*_adj_<0.05 (data not shown).

Taken together, type 1 and type 17 responses are more pronounced in colitis tissue of non-responders, but we found no evidence supporting that this difference is driven by mechanistical involvement of Th17 cells, in particular pathogenic Th17.1 cells, in steroid-unresponsive ICI colitis.

## Discussion

Current irAE treatment strategies relying on empirical use of high-dose steroids in first-line and targeted immunosuppression for steroid-refractory cases are suboptimal, since both high-dose steroids and second-line immunosuppressants for irAEs have been associated with worse survival. However, use of alternative strategies that can limit cumulative immunosuppression exposure, including early use of targeted immunosuppressants, and optimal candidate selection for such approach is limited by insufficient knowledge of irAE pathogenesis, including poor understanding of steroid resistance mechanisms. In the present study, we took a multi-omics approach using blood and tissue from patients with mostly gastro-intestinal irAEs, to identify pre-steroids predictors of response and characterize immunological patterns associated with steroid non-response. In summary, steroid non-response was reflected in blood by trends for increased Tc1-like CD8^+^ T cells and serum IL-17, IL-6, IL-12 and IL-23 before, and persistent T cell activation and proliferation after initiation of steroids. In colitis tissue, an enhanced mixed type 1/type 17 immune response showed the strongest association with steroid non-responsive versus steroid-responsive colitis. While activated CD4^+^ memory T cells were enriched in colitis tissue of non-responders, detailed analysis of tissue CD4^+^ T cells in a larger cohort did not support the involvement of specific CD4^+^ T cell subsets in steroid-unresponsive irAEs.

We and others have previously shown that mixed type 1/type 17 responses are dominant in irAEs,^9,25-27^ with type 17 skewing particularly observed after ipilimumab-containing therapy.^9,27^ Consistent with previous reports,^27,28^ we observed elevated IL-17 (as well as IL-6 and IL-17-inducible MMP-3) in serum of non-responders prior to steroid initiation. Furthermore, a type 17 signature was enriched in transcriptomic data from bulk tissue of ICI colitis patients not responding to steroids. These findings are concordant with small studies on blood of ICI arthritis patients suggesting a role for Th17/Tc17 cells in steroid resistance.^27,28^ Increased type 17 skewing could simply reflect broader immune activation in the context of severe irAEs. However, because Th17-pathway-directed treatment strategies have already shown promise in treating irAEs while enhancing anti-tumor responses,^25,29,30^ we explored cues to support whether these therapies could also abrogate steroid resistance in irAEs. Tissue abundance of *IL17A-* and *IL22*-expressing CD4^+^ clusters was not associated with steroid response. We may have underestimated the abundance of Th17/Th22 cells focusing on *IL17A*^+^ and *IL22*^+^ clusters, since expression of secreted proteins including cytokines is only moderately reflected on the transcriptomic level.^31^ In addition, the entire pool of Th17/Th22 cells may actually represent subsets with different inflammatory potential. Th17 cells, and Th17.1 cells in particular, have been implicated in steroid resistance in various autoimmune diseases, including IBD, Th17-high asthma and multiple sclerosis.^24,32,33^ Th17.1 cells feature a pathogenic phenotype (producing both IFN-γ and IL-17) and are specifically associated with *ABCB1*, or multi-drug resistant type 1 (*MDR1*) expression, which actively expels prednisolone and dexamethasone from the cytoplasm.^34,35^ Enrichment analysis in colitis tissue of a Th17.1 signature, comprising *ABCB1*, did not reveal an association with steroid response in our cohort either. Taken together, we were unable to demonstrate a role for Th17 cells as drivers of steroid non-response in irAEs, specifically ICI colitis. Other sources of IL-17, such as type 3 innate lymphoid cells, could alternatively be responsible for the type 17 signature enrichment and increased serum IL-17 in non-responders.

Although limited by the observational nature of our data, we showed that lymphocyte infiltration was lower in tissue of steroid-responding patients within 24h after initiation of steroids compared to pre-steroids biopsies. Confounding by severity is unlikely, as patients undergoing endoscopy after initiation of steroids on average had higher grade colitis than patients examined pre-steroids. Molecular analysis of mucosal inflammation across timepoints was underpowered, but showed the same trend of lower inflammation in steroid-responders within 24h of initiation of steroids. Together, these data suggest very early immunological effects of steroids in steroid-responding patients with ICI colitis. As genomic effects of steroids take more than 24h to come into effect, lower lymphocyte (but not neutrophil) infiltration could be explained by lymphotoxicity through membrane-bound glucocorticoid receptors or other non-genomic mechanisms.^36^ Besides steroid-induced apoptosis, peaking after 72h and primarily affecting activated T cells based on in vitro studies,^37,38^ redistribution of lymphocytes could also explain their lower tissue abundance. Importantly, molecular indices of mucosal inflammation have not been assessed sooner than 4 weeks after treatment initiation in IBD trials.^17^

We also quantitively assessed lymphocyte and neutrophil infiltration in tissue slides of ICI colitis, but found no association with response to steroids. The extent of histologic and endoscopic inflammation, particularly ulceration, has been associated with need for second-line immunosuppression in ICI colitis.^39-43^ Histological inflammation measured by the Robarts Histopathology Index (RHI), but not the pattern of inflammation (acute, chronic-active or microscopic), was associated with biological use in multivariate analysis of ICI colitis.^44^ In contrast, chronic histological abnormalities, such as basal plasmacytosis and crypt architectural distortion, independently predict relapse in ulcerative colitis.^45,46^ The limited amount of data in ICI colitis may thus indicate that histological indices of chronicity in ICI colitis do not necessarily reflect the necessity for treatment escalation. In support of this, time-to-irAE-onset since ICI initiation was not associated with steroid response rates for most irAEs including colitis,^47^ although time between symptom onset and treatment initiation could constitute a better proxy for chronicity. Neutrophil infiltration is an important criterion in the RHI, worth up to 15 (of maximum 33) points allocated for lamina propria and epithelial neutrophil infiltration. In our study, neither pre-steroids neutrophil, nor lymphocyte infiltration predicted steroid non-response. This may be due to our quantitative approach, lacking qualitive detail on the extent of inflammation and leukocyte distribution patterns. These findings underscore that in standardizing histological evaluation of ICI colitis,^48^ qualitative aspects of inflammation may hold more prognostic value than purely quantitative criteria.

We are aware that our study has limitations. Despite the large number of prospectively enrolled patients in this hypothesis-generating study, the number of patients with irAEs and complete blood sampling was too limited to consider tumor response in addition to ICI type as covariates. Moreover, not all tissue could be obtained before initiation of steroids, although this enabled description of possible mucosal effects early after initiation of steroids.

In conclusion, a trend for expansion of a mixed type 1/type 17 immune response in blood of patients with irAEs who are not responding to steroids was reflected by enrichment of especially type 1 and type 17 signatures in ICI colitis tissue, but could not be consolidated by detailed T-helper subset analysis. We provide preliminary molecular and histological evidence that steroids may sort effects in tissue shortly after treatment initiation in steroid-responding patients with irAEs. Furthermore, our translational data may aid in constructing standardized histological evaluation criteria for ICI colitis and inform the design of studies to investigate new treatment strategies for irAEs.

## Methods

### Patient selection, sample collection and outcome definition

Peripheral blood mononuclear cell (PBMC), serum and snap-frozen colon biopsies from patients enrolled in the UNICIT biobank study were included. In this biobank study, adult patients undergoing immune checkpoint inhibitor treatment for solid malignancies at University Medical Center Utrecht are enrolled and blood and stool samples are collected early during treatment, upon onset of irAEs and after each line of immunosuppressive treatment for irAEs, together with biopsies in case of diagnostic procedures. Details of the UNICIT cohort have been described previously.^9^

For the present study, PBMC samples were selected for flow cytometry from UNICIT patients that developed clinically relevant irAEs, defined as CTCAE grade ≥2 and requiring ≥0.5 mg/kg/day corticosteroids. Moreover, the selection was enriched for melanoma (75%) and gastro-intestinal toxicity (75%), with paired samples before and after initiation of systemic steroids available (**Supplementary Table 1**). The extended serum cohort consisted of serum samples from all patients in the flow cytometry cohort, supplemented with extra pre-steroids-only samples from UNICIT patients treated with systemic steroids for irAEs, again enriched for melanoma (77.5%) and gastro-intestinal toxicity (80.0%; **Supplementary Table 2**). Patients with simultaneous gastro-intestinal infections were excluded. For bulk RNA-sequencing, all snap-frozen colon biopsies from UNICIT patients with histologically confirmed ICI colitis who required immunosuppressive treatment were selected (**Supplementary Table 3**). Digital images of hematoxylin & eosin (H&E) colon tissue slides from patients included for bulk RNA-seq were also used, along with H&E images from patients diagnosed with ICI colitis selected from a retrospective registry containing patients with ICI colitis treated at University Medical Center Utrecht until January 2023 (**Supplementary Table 4**). Healthy donor blood (for PBMCs) was obtained from the University Medical Center Utrecht Mini Donor Service.

Blood was collected in sodium heparin tubes for PBMCs and clot-blood tubes for serum. PBMCs were isolated with Ficoll-Paque density-gradient centrifugation (GE Healthcare), frozen in RPMI-1640 medium (Gibco) with 2 mM L-glutamine (Gibco), 100 IU/ml penicillin/streptomycin (Gibco), 20% fetal bovine serum (FBS; Invitrogen) and 10% dimethylsulfoxide and stored at −196 °C. Serum was isolated and frozen at −80 °C within 4h of blood collection. Colon biopsies in the RNA-seq cohort and retrospective registry were obtained during flexible sigmoidoscopy (81%) or full colonoscopy (19%) with pinch biopsy forceps from macroscopically inflamed mucosa, or at random if no macroscopic abnormalities were present. Biopsies for RNA-seq were snap-frozen in liquid nitrogen immediately after collection and stored at −80 °C.

Steroid non-responders were defined as patients requiring any second-line immunosuppression to achieve complete remission of irAEs before steroids had been fully tapered.

### Spectral flow cytometry

Cells were thawed and plated at ∼1.0-3.0×10^6^ per well for spectral flow cytometry. Cells were first restimulated with 20 ng/ml phorbol 12-myristate 13-acetate (PMA; Sigma-Aldrich) and 1.0 μg/ml ionomycin (Sigma-Aldrich; 4h, 37°C, 5% CO_2_), with GolgiStop (0.26% monensin; BD Biosciences, 1:1,500) added for the last 3.5h. Cells were stained with fixable viability dye eFluor506 (Invitrogen; 30 min, 4°C), washed and then stained with the surface mix (**Supplementary Table 5**) in FACS buffer (phosphate buffered saline [PBS, Sigma-Aldrich], 2% FBS, 0.1% sodium azide; 25 min, 4°C). Cells were fixed and permeabilized with the eBioscience FoxP3/transcription factor fixation/permeabilization kit (Invitrogen) for 30 min at 4°C, followed by the intracellular/intranuclear mix (**Supplementary Table 5**) in permeabilization buffer (30 min, 4°C). Cells were kept in the dark at 4°C until the next day and were then measured on a Cytek® Aurora™ 5L spectral flow cytometer. Cytek® FSP™ CompBeads were used for all single-stain controls, except for CD8, CD45RA and TNF-α, which were prepared with cells. Unstained controls were prepared with cells from each individual measured. Bridging controls from one patient and one healthy donor were taken along in all three batches. Before measurement, daily quality control was performed with SpectroFlo QC beads. Samples were randomized over different batches by response to steroids (primary outcome) and longitudinal samples from the same subject were included in the same batch and measured consecutively.

After spectral unmixing with SpectroFlo software (Cytek), FlowJo (Tree Star) was used to pre-gate live singlets (**Supplementary Figure 4**). Further analysis was exclusively performed in R and based on a published pipeline.^49^ First, data were archsinh-transformed with cofactor=6000 for all channels except FSC/SSC and quality control was performed with the *peacoQC* package (IT_limit=0.55, MAD=6).^50^ After pre-gating and quality control, over 22 million cells remained for further analysis. Batch effect and intra-patient variation not attributable to biology was corrected by a custom landmark-based normalization method similar to fdaNorm,^51^ but using the downslope infliction point instead of density maxima as landmarks.

Next, data were clustered using a combination of *FlowSOM* and *Seurat*. To this end, data were first centered log ratio (CLR) normalized and then clustered with *FlowSOM* (nClus=50)^52^ based on CD57, CD56, FoxP3, CD161 (KLRB1), CD39, CD3, CD16, CCR7, HLA-DR, CD4, CD8, CD14 and CD45RA to identify immune cell subsets present within the PBMC fraction. Subsequently, we visualized cell clustering in two-dimensional space using *Seurat*.^23^ To this end, we randomly drew a 1% subset (∼220,000 cells) of the complete data set. Data were CLR-normalized and scaled with *ScaleData()*, followed by principal component analysis (PCA), Uniform Manifold Approximation and Projection (UMAP) and clustering with *FindNeighbors()* based on FSC/SSC along with the same markers used for the Self-Organizing Map (FlowSOM). Optimal clustering resolution was assessed with *Clustree*.^53^ Major subsets were identified by grouping together similar FlowSOM-clusters (**Supplementary Figure 1a**). FlowSOM metacluster 1 (containing 0.018% of all cells) was discarded, because it contained outlier events with 21 (biologically unrelated) markers showing extremely high expression compared to other metaclusters, suspect for residual debris. Next, abundance of cell subsets and their functional profiles was assessed by analyzing expression levels of cytokines, receptors and transcription factors.

### PD-1 receptor occupancy assay

T cell fractions completely or partially bound by PD-1-blocking therapeutic antibodies were estimated using a previously described receptor occupancy assay.^54^ Briefly, in this approach cells are stained both with anti-PD-1 clone EH12.1, competitive with nivolumab and pembrolizumab, and with anti-human IgG4 (specifically binding nivolumab or pembrolizumab). For this assay, 1.0×10^6^ cells were plated and stained, fixed and permeabilized similarly to the spectral flow cytometry protocol, but now after 5h incubation (37°C, 5% CO_2_) in presence of GolgiStop (1:1,500), without PMA/ionomycin, to enable measurement of CXCL13,^55^ and using the staining mix in **Supplementary Table 6**. Cells were measured the next day on an LSR Fortessa. Data were analyzed in FlowJo according to the gating strategy in **Supplementary Figure 4**.

### ICI-bound PD-1 internalization assay

Pre-steroids PBMCs from 4 patients (2 with highest CD4^+^ T cell PD-1 RO and 2 with lowest CD4^+^ T cell PD-1 RO) were stained with the staining mix in **Supplementary Table 7** as described above, but this time immediately after thawing. Each patient sample was then split in three and each aliquot was measured on an ImageStream X Mark II at 0, 30 and 60 min after completion of staining, to assess internalization of ICI-bound PD-1 receptor over time in the ex vivo situation. Internalization was quantified by the internalization feature, derived from the ratio of intracellular-to-total-cell intensity of PE (IgG4), using the Internalization Wizard in IDEAS software. The standard deviation of the Internalization feature was approximated by 1.25 × MAD (median absolute deviation).

### Serum multiplex immunoassay

Serum concentrations of 26 analytes (IL-5, IL-6, IL-10, IL-12, IL-13, IL-17, IL-21, IL-23, TNF-α, IFN-γ, APRIL, CCL2, CCL4, CCL17 [TARC], CXCL9, CXCL10, CXCL13, CD40L, soluble IL-2Rα, granzyme B, TGF-β1, TACI, galectin-9 [Gal9], MMP-3, MMP-9 and MMP-10) were measured using an in-house developed and validated (ISO9001 certified) multiplex immunoassay (Center for Translational Immunology, University Medical Center Utrecht) based on Luminex technology (xMAP, Luminex Austin TX USA). The assay was performed as described previously.^56^ A-specific heterophilic immunoglobulins were preabsorbed from all samples with heteroblock (Omega Biologicals, Bozeman MT, USA). Acquisition was performed with the Biorad FlexMAP3D (Biorad laboratories, Hercules USA) in combination with xPONENT software version 4.2 (Luminex). Data was analyzed by 5-parametric curve fitting using Bio-Plex Manager software, version 6.1.1 (Biorad).

### Bulk RNA-sequencing

#### RNA isolation

RNA isolation was performed using the RNeasy Mini kit (Qiagen). First, 2 mL microcentrifuge tubes containing a 5mm stainless steel bead (Qiagen) were precooled on dry ice, while the adapter of the TissueLyser LT (Qiagen) was placed on regular ice. After 15 min, frozen biopsies were transferred into the 2 mL tubes and placed in the adapter, 600 µL Lysis buffer per sample (containing 10 µL β-mercaptoethanol per mL RLT buffer) was added and the TissueLyser was switched on as quickly as possible for disruption and homogenization (total 6 cycles × 1 min, 25 Hz). In between cycles, the tissue adapter containing the samples was cooled on regular ice for 30 sec. Next, tubes were briefly centrifuged to remove foam and debris from the lids (5 sec., 13,000 rpm, RT). The lysates were pipetted into new Eppendorf tubes and centrifuged (3 min, 13,000 rpm, RT). The supernatant was transferred into new 1.5 mL tubes without debris or pellet, to which 0.5× volumes of 70% ethanol were added and then lysates were transferred to the RNeasy spin columns. Next steps of the RNeasy Mini kit were performed according to the manufacturer’s protocol. This homogenization and RNA isolation protocol yielded optimal and balanced RNA quantity, purity and quality (measured by an Implen N60/N50 NanoPhotometer and Agilent 2100 Bioanalyzer system using the Agilent RNA 6000 Pico Kit).

#### RNA sequencing

Library preparation was performed at GenomeScan with the NEBNext Ultra II Directional RNA Library Prep Kit (Illumina) according to protocol NEB #E7760S/L) and included mRNA isolation by oligo-dT magnetic beads. Sufficient quality and yield for all samples before sequencing was verified on a Fragment Analyzer (Agilent). Twenty-million paired-end (150 bp) reads per sample were sequenced on a NovaSeq 6000 sequencer. Reads were then trimmed with *fastp* v0.23.5, mapped to a human reference genome (GRCh38.p13) based on Burrows-Wheeler Transform (STAR2 v2.7.10) and count tables were created with *HTSeq* v2.0.2. Transcripts-per-million (TPM) values were calculated with *Cufflinks* v2.2.1.

#### Bulk RNA-seq analysis

Low-abundant genes (total ≤ 5 counts or ≤ 0.01 TPM across samples) were filtered. Absolute abundance estimates of 22 immune cell subsets were deconvoluted with the CIBERSORT algorithm in absolute mode using the LM22 signature matrix and untransformed TPM files.^16^ Only samples deconvoluted at *P*_perm_<0.05 were kept for further analysis. We additionally leveraged the LM7 signature mix in an attempt to obtain more reliable estimations of γδ T cells,^57^ given their high expected abundance in colonic mucosa. However, LM7 and LM22 results with respect to γδ T cells were similar and therefore, we exclusively used LM22 results. Gene set variation analysis (GSVA) was performed with *GSVA* v1.52.3,^58^ using logTPM values. We measured the bMIS_UC gene set (**Supplementary Table 8**), validated in ulcerative colitis to molecularly assess mucosal inflammation and predict treatment outcome.^17^ In addition, a published Th17.1 gene signature^24^ and custom gene sets were analyzed, deliberately containing mutually exclusive Th1-, Th2- and Th17-related genes (**Supplementary Table 8**) to maximize any signal for immune skewing. A core signature for mature B cells and plasma cells was compiled based on human scRNA-seq results.^59^

### Pooled single-cell transcriptomic data meta-analysis

CD4^+^ T cell transcriptomic data from 37 patients with ICI colitis (70.3% steroid-naïve upon endoscopy) for whom response to steroids was reported from three single-cell RNA-sequencing studies were reanalyzed with *Seurat*.^20-22^ Non-responders were defined as patients who were reported to have received second-line immunosuppressive therapy after steroids.

From the Thomas et al. dataset (GEO: GSE206301),^21^ patients of interest were selected from CD4^+^ T cell transcriptomic data provided in H5ad format. Separate Seurat objects were created for individual patients, only cells with 200-3,000 features and <10% mitochondrial RNA were kept and TCR genes were silenced.^60^ The dataset was split by patient, *SCTransform()* with *glmGamPoi* was run on individual Seurat objects and then data from all patients were merged again. From the Gupta et al. dataset (GEO: GSE189040),^20^ patients of interest could be directly selected from a Seurat object containing clustered and annotated T cell populations, from which CD8^+^ T cell subsets were excluded. From the Luoma et al. dataset (GEO: GSE144469),^22^ CD3^+^ sorted transcriptomic data were transformed as Seurat object and subjected to quality control and *SCTransform()* as outlined above. Variable features (n=3,000) were selected with *SelectIntegrationFeatures()*, followed by dimensionality reduction (*RunPCA(), RunUMAP()* with the first 30 dimensions) and clustering (*FindNeighbors()* with first 30 dimensions and *FindClusters()* at resolution=1.0) to obtain *CD4*-expressing clusters, which were selected for further analysis.

Next, only features detected in all three datasets were kept and all datasets were merged and clustered as described above. Residual CD8^+^ T cell clusters were dropped, cells were clustered again and mapped as unintegrated UMAP plot to evaluate the extent of study-derived batch effect. The original three datasets were then merged again, subjected to *SCTransform()* and PCA, followed by *IntegrateLayers()* using SCT-transformed data and reciprocal PCA (RPCA).^23^ Residual CD8^+^ T cell clusters were dropped again and dimensionality reduction and clustering was repeated using integrated dimensions. Cluster-defining markers were identified with *FindAllMarkers()* and cell fractions per CD4^+^ T cell cluster were calculated for all patients. Clusters were not individually annotated, but Th17 and Th22 clusters were specifically identified based on *IL17A* and *IL22* expression. GSVA was performed on pseudobulk data generated with *AverageExpression()* using SCT data. Differential gene expression was performed with *DESeq2* using pseudobulk data from raw RNA counts, adjusting for GSE dataset of origin.^61^

### Automatic nucleus segmentation and cell type classification in colitis H&E slides

Whole slide images of hematotoxylin & eosin (H&E) stained colon specimens were analyzed using the Hover-NeXt model trained on the Lizard dataset.^18,62^ This model, trained with nuclei annotations from normal colon tissue and colorectal cancer, segments and classifies all visible nuclei into connective tissue nuclei, epithelial nuclei, neutrophils, eosinophils, plasma cells or lymphocytes. Next, tissue containing areas were manually annotated using QuPath.^63^ These tissue annotations were used to filter out false positive nuclei annotations outside tissue area using the GeoPandas package. This resulted in a .GEOjson file with coordinates and classifications of nuclei inside the tissue area which was used for further analysis.^64^

### Statistics

Median follow-up time was estimated with the reverse Kaplan-Meier method using the *survival* package. Unless otherwise specified, continuous variables between two or more than two groups were analyzed by Wilcoxon or Kruskal-Wallis tests without formal post-hoc test, respectively. Correlations were analyzed by Spearman’s correlation. *P* values were adjusted for multiple testing with Benjamini-Hochberg procedures. Unadjusted *P* values were also reported given increased type II error rates with multiple-testing correction, to address the hypothesis-generating nature of this study. Linear regression models were fit to quantify the relation of cell subsets with steroid response adjusted for ICI treatment type and *vice versa* with the *stats* package. Trends in cross-sectional data across different timepoints were visualized with local regression using locally estimated scatterplot smoothing (LOESS) embedded in *geom_smooth()* within *ggplot2*, with timepoints beyond 7 days winsorized.

### Ethics

The UNICIT biobank study was not considered subject to the Dutch Medical Research with Human Subjects Law by the medical research ethics committee of the University Medical Center Utrecht. The UMC Utrecht biobank review committee approved the UNICIT biobank protocol (TCbio 18-123) and granted permission for use of human biospecimens for this study (TCbio 23-200). All participants in this biobank study provided written informed consent in line with de Declaration of Helsinki. Informed consent was waived for the use of data from patients who had not opted-out for use of their health data for research purposes and who had been included in a single-center retrospective colitis registry.

## Data Availability

All data produced in the present study are available upon reasonable request to the authors, to the extent permitted by applicable law.

## Acknowledgements

We thank the patients, their families and caregivers and healthy donors for their willingness to participate in this study; clinical staff and UNICIT consortium members for contributing to patient accrual, sample collection and biobanking; and the Multiplex Core Facility of the University Medical Center Utrecht for performing the multiplex immunoassays. Figure 1 and icons used in Figures 2-4 have been created with www.biorender.com.

## Funding

This investigator-initiated study received funding from Bristol Myers Squibb (grant number CA209-6JY), paid to institution.

## Conflict of interest

MJMvE: none. MMvdW: none. HBK: none. NMMD: none. MS: none. RJV: none. FDMvS: none. BO: none. SN: none. KPMS: Consulting/advisory relationship: Abbvie, Sairopa. Research funding: TigaTx, Bristol Myers Squibb, Philips, Genmab, Pierre Fabre. Honoraria: Bristol Myers Squibb. All paid to institution. FW: has advisory relationships with Janssen and Takeda, and received research funding from Takeda, Galapagos, BMS, Sanofi, and Leo Pharma.

## Notes

### Author Declarations

The biobank review committee of the University Medical Center Utrecht gave ethical approval for this work (Biobank protocol TC-bio 18-123 and Material release protocol TC-bio 23-200).

