## Supplementary material for "Blood and tissue correlates of steroid non-response in checkpoint inhibition-induced immune-related adverse events"

### Supplementary Tables

#### Supplementary Table 1 Flow cytometry PBMCs cohort

| | | Steroid responder (N=13) | Steroid non-responder (N=11) | Healthy donor (N=6) |
|---|---|---|---|---|
| **Female sex, n (%)** | | 4 (30.8%) | 6 (54.5%) | 4 (66.7%) |
| **Median age, yr (IQR)** | | 61 (58-70) | 67 (59-70) | 62 (61-63) |
| **Tumor type, n (%)** | *melanoma* | 9 (69.2%) | 9 (81.8%) | N/A |
| | *non-small cell lung cancer* | 1 (7.7%) | 2 (18.2%) | |
| | *renal cell carcinoma* | 3 (23.1%) | 0 (0.0%) | |
| **Tumor stage IV, n (%)** | | 12 (92.3%) | 8 (72.7%) | N/A |
| **ICI Treatment, n (%)** | *ipilimumab monotherapy* | 0 (0.0%) | 1 (9.1%) | N/A |
| | *ipilimumab + nivolumab* | 10 (76.9%) | 6 (54.5%) | |
| | *anti-PD-(L)1 monotherapy* | 3 (23.1%) | 4 (36.4%) | |
| **Main irAE type, n (%)** | *colitis* | 5 (38.5%) | 8 (72.7%) | N/A |
| | *gastritis/duodenitis* | 3 (23.1%) | 2 (18.2%) | |
| | *hepatitis* | 3 (23.1%) | 1 (9.1%) | |
| | *pneumonitis* | 1 (7.7%) | 0 (0.0%) | |
| | *encephalitis* | 1 (7.7%) | 0 (0.0%) | |
| **Main irAE CTCAE grade, n (%)** | *II* | 4 (30.8%) | 3 (27.3%) | N/A |
| | *III* | 9 (69.2%) | (63.6%) | |
| | *IV* | 0 (0.0%) | 1 (9.1%) | |
| **≥1 concomittant irAE(s), n (%)** | | 5 (38.5%) | 5 (45.5%) | N/A |
| **Median time between pre- and post-steroids sample, days (IQR)** | | 15 (9-28) | 6 (5-12) | N/A |

### Supplementary Table 2 Extended serum cohort

| | | Steroid responder (N=14) | Steroid non-responder (N=17) |
|---|---|---|---|
| **Female sex, n (%)** | | 7 (50.0%) | 9 (52.9%) |
| **Median age, yr (IQR)** | | 69 (60-72) | 58 (52-71) |
| **Tumor type, n (%)** | *melanoma* | 8 (57.1%) | 14 (82.4%) |
| | *non-small cell lung cancer* | 1 (7.1%) | 1 (5.9%) |
| | *squamous cell carcinoma* | 0 (0.0%) | 1 (5.9%) |
| | *urothelial cell carcinoma* | 1 (7.1%) | 0 (0.0%) |
| | *renal cell carcinoma* | 4 (28.6%) | 0 (0.0%) |
| | *melanoma, NSCLC, mesothelioma* | 0 (0.0%) | 1 (5.9%) |
| **Irresectable tumor stage III and IV, n (%)** | | 14 (100.0%) | 16 (94.1%) |
| **ICI Treatment, n (%)** | *ipilimumab + nivolumab* | 12 (85.7%) | 13 (76.5%) |
| | *anti-PD-1 monotherapy* | 1 (7.1%) | 2 (11.8%) |
| | *anti-PD-1 + targeted/chemotherapy* | 1 (7.1%) | 2 (11.8%) |
| **Main irAE type, n (%)** | *colitis* | 8 (57.1%) | 12 (70.6%) |
| | *gastritis/duodenitis* | 2 (14.3%) | 4 (23.5%) |
| | *hepatitis* | 4 (28.6%) | 0 (0.0%) |
| | *ileitis terminalis* | 0 (0.0%) | 1 (5.9%) |
| **Main irAE CTCAE grade, n (%)** | *II* | 8 (57.1%) | 8 (47.1%) |
| | *III* | 6 (42.9%) | 9 (52.9%) |
| **≥1 concomittant irAE(s), n (%)** | | 6 (42.9%) | 7 (41.2%) |

### Supplementary Table 3 Bulk RNA-seq colon biopsy cohort

| | | Steroid responder (N=10) | Steroid non-responder (N=14) |
|---|---|---|---|
| **Female sex, n (%)** | | 2 (20.0%) | 7 (50.0%) |
| **Median age, yr (IQR)** | | 67 (63-71) | 60 (49-75) |
| **Tumor type, n (%)** | *melanoma* | 7 (70.0%) | 12 (85.7%) |
| | *urothelial cell carcinoma* | 1 (10.0%) | 1 (7.1%) |
| | *colorectal carcinoma* | 0 (0.0%) | 0 (0.0%) |
| | *non-small cell lung cancer* | 0 (0.0%) | 1 (7.1%) |
| | *renal cell carcinoma* | 2 (20.0%) | 0 (0.0%) |
| **Tumor stage IV, n (%)** | | 8 (80.0%) | 14 (100.0%) |
| **ICI Treatment, n (%)** | *ipilimumab monotherapy* | 0 (0.0%) | 1 (7.1%) |
| | *ipilimumab + nivolumab* | 4 (40.0%) | 9 (64.3%) |
| | *anti-PD-(L)1 monotherapy* | 6 (60.0%) | 3 (21.4%) |
| | *anti-PD-1 + targeted/chemotherapy* | 0 (0.0%) | 1 (7.1%) |
| **CTCAE grade colitis, n (%)** | *I* | 1 (10.0%) | 1 (7.1%) |
| | *II* | 6 (60.0%) | 6 (42.9%) |
| | *III* | 3 (30.0%) | 7 (50.0%) |
| **Mayo endoscopic score, n (%)** | *0* | 1 (10.0%) | 1 (7.1%) |
| | *1* | 7 (70.0%) | 8 (57.1%) |
| | *2* | 1 (10.0%) | 4 (28.6%) |
| | *3* | 1 (10.0%) | 1 (7.1%) |
| **No steroids at endoscopy, n (%)** | | 6 (60.0%) | 5 (35.7%) |
| **Median steroid duration at endoscopy, days (IQR)** | | 0 (0-1) | 1 (0-6) |

### Supplementary Table 4 Extended H&E colon biopsy cohort

| | | Steroid responder (N=25) | Steroid non-responder (N=30) |
|---|---|---|---|
| **Female sex, n (%)** | | 9 (36.0%) | 19 (63.3%) |
| **Median age, yr (IQR)** | | 66 (54-71) | 69 (60-75) |
| **Tumor type, n (%)** | *melanoma* | 10 (40.0%) | 22 (73.3%) |
| | *urothelial cell carcinoma* | 3 (12.0%) | 1 (3.3%) |
| | *head/neck cancer* | 2 (8.0%) | 1 (3.3%) |
| | *hepatocellular carcinoma* | 2 (8.0%) | 0 (0.0%) |
| | *non-small cell lung cancer* | 5 (20.0%) | 3 (10.0%) |
| | *renal cell carcinoma* | 3 (12.0%) | 3 (10.0%) |
| **Tumor stage IV, n (%)** | | 22 (88.0%) | 24 (80.0%) |
| **ICI Treatment, n (%)** | *ipilimumab monotherapy* | 1 (4.0%) | 4 (13.3%) |
| | *ipilimumab + nivolumab* | 9 (36.0%) | 13 (43.3%) |
| | *anti-PD-(L)1 monotherapy* | 14 (56.0%) | 10 (33.3%) |
| | *anti-PD-1 + targeted/chemotherapy* | 1 (4.0%) | 3 (10.0%) |
| **CTCAE grade colitis, n (%)** | *I* | 1 (4.0%) | 1 (3.3%) |
| | *II* | 15 (60.0%) | 10 (33.3%) |
| | *III* | 9 (36.0%) | 17 (56.7%) |
| | *IV* | 0 (0.0%) | 1 (3.3%) |
| | *V* | 0 (0.0%) | 1 (3.3%) |
| **No steroids at endoscopy, n (%)** | | 10 (40.0%) | 15 (50.0%) |
| **Median steroid duration at endoscopy, days (IQR)** | | 0 (0-1) | 1 (0-2) |

### Supplementary Table 5 Spectral flow cytometry panel

| Detector | Fluorochrome | Target | Clone | Reactivity | Dilution | Staining | Company | Catalogus no. | Lot no. |
|---|---|---|---|---|---|---|---|---|---|
| UV2 | Spark UV387 | CD45RA | HI100 | anti-human | 50 | Surface | Biolegend | 304179 | B389525 |
| UV7 | BUV496 | CD3 | UCHT1 | anti-human | 50 | Surface | BD | 612940 | 3109436 |
| UV9 | BUV563 | CD38 | HB7 | anti-human | 100 | Surface | BD | 741446 | 3300104 |
| UV11 | BUV661 | CD134 | ACT35 | anti-human | 50 | Surface | BD | 750645 | 3171474 |
| UV14 | BUV737 | TNFa | MAb11 | anti-human | 100 | Intracullular | Invitrogen | 367-7349-42 | 2778202 |
| V1 | BV421 | TCF7 | S33-9 | anti-human/mouse | 200 | Intracullular | BD | 566692 | 2129120 |
| V2 | Super Bright 436 | beta1 | TS2/16 | anti-human | 50 | Surface | ThermoFisher | 15923374 | 2547810 |
| V3 | Pacific Blue | GzmB | GB11 | anti-human | 100 | Intracullular | Biolegend | 515407 | B358356 |
| V5 | BV480 | beta7 | FIB504 | anti-human/mouse | 400 | Surface | BD | 746792 | 3171350 |
| V7 | eFluor506 | *Fixable viability dye* | N/A | N/A | 2000 | N/A | eBioscience | 65-0866-14 | 2526187 |
| V8 | BV570 | CD16 | 3G8 | anti-human | 200 | Surface | Biolegend | 302035 | B379014 |
| V10 | BV605 | CCR7 | G043H7 | anti-human | 25 | Surface | Biolegend | 353223 | B381861 |
| V11 | BV650 | CD25 | BC96 | anti-human | 100 | Surface | Biolegend | 302633 | B370849 |
| V13 | BV711 | IL-17A | BL168 | anti-human | 100 | Intracullular | Biolegend | 512327 | B359414 |
| V14 | BV750 | PD-1 | EH12.1 | anti-human | 25 | Surface | BD | 747446 | 3300142 |
| V15 | BV785 | CXCR3 | G025H7 | anti-human | 25 | Surface | Biolegend | 353737 | B384611 |
| B2 | Alexa Fluor 488 | FoxP3 | 259D | anti-human | 20 | Intracullular | Biolegend | 320211 | B358133 |
| B3 | Spark blue 550 | CD8 | SK1 | anti-human | 200 | Surface | Biolegend | 344759 | B361158 |
| B4 | Spark Blue 574 | CD14 | HCD14 | anti-human | 25 | Surface | Biolegend | 325635 | B376662 |
| B8 | PerCP | HLA-DR | L243 | anti-human | 100 | Surface | BD | 347402 | 2265916 |
| B9 | PerCP-Cy5.5 | IL-13 | JES10-5A2 | anti-human | 50 | Intracullular | Biolegend | 501912 | B382906 |
| B9 | BB700 | CD39 | TU66 | anti-human | 100 | Surface | BD | 745904 | 3171339 |
| B10 | PerCP-eFluor710 | CTLA-4 | 14D3 | anti-human | 100 | Intracullular | ThermoFisher | 46-1529-41 | 2338619 |
| B14 | PerCP-Fire806 | CD4 | SK3 | anti-human | 400 | Surface | Biolegend | 344693 | B391368 |
| R1 | APC | CD57 | HNK-1 | anti-human | 100 | Surface | Biolegend | 359609 | B397031 |
| R3 | Spark NIR685 | IFNy | 4S.B3 | anti-human | 50 | Intracullular | Biolegend | 502551 | B314651 |
| R4 | Alexa Fluor 700 | CD161 | HP-3G10 | anti-human | 50 | Surface | Biolegend | 339941 | B388780 |
| R7 | APC-Fire750 | CD56 | QA17A16 | anti-human | 400 | Surface | Biolegend | 985904 | B402872 |
| YG1 | PE | IL-10 | JES3-19F1 | anti-human | 20 | Intracullular | BD | 554706 | 335763 |
| YG1 | RY586 | IL-5 | TRFK5 | anti-human/mouse | 200 | Intracullular | BD | 568530 | 2222943 |
| YG3 | PE-Dazzle594 | LAG-3 | 11C3C65 | anti-human | 50 | Surface | Biolegend | 369331 | B379163 |
| YG4 | PE-Fire640 | CD69 | FN50 | anti-human | 50 | Surface | Biolegend | 310959 | B364692 |
| YG5 | PE-Cy5 | TIM-3 | F38-2E2 | anti-human | 200 | Surface | Sony | 2325255 | 284479 |
| YG9 | PE-Cy7 | Ki67 | Ki-67 | anti-human | 800 | Intracullular | Biolegend | 350525 | B361326 |
| YG10 | PE-Fire810 | CCR4 | L291H4 | anti-human | 200 | Surface | Biolegend | 359433 | B387376 |

### Supplementary Table 6 Conventional flow cytometry panel

**Primary mix**

| Fluorochrome | Target | Clone | Reactivity | Dilution | Staining | Company | Catalogus no. | Lot no. |
|---|---|---|---|---|---|---|---|---|
| FITC | Ki67 | MIB-1 | anti-human | 200 | Intracellular | DAKO | F7268 | 41365106 |
| PerCP-Cy5.5 | Integrin beta7 | FIB27 | anti-human | 50 | Surface | Biolegend | 121008 | B387527 |
| APC | CXCL13 | 53610 | anti-human | 50 | Intracellular | R&D | IC801A | ABAG0419091 |
| AF700 | CD3 | UCHT1 | anti-human | 50 | Surface | Biolegend | 300424 | B399038 |
| APC-Fire750 | CCR7 | G043H7 | anti-human | 12,5 | Surface | Biolegend | 353246 | B338294 |
| PB | CD45RA | HI100 | anti-human | 100 | Surface | Biolegend | 304117 + 304118 | B397330 |
| eFluor506 | *Fixable viability dye* | N/A | N/A | 2000 | N/A | eBioscience | 65-0866-14 | 2526187 |
| BV711 | PD-1 | EH12.1 | anti-human | 100 | Surface | BD | 564017 | 2076556 |
| BV785 | CD4 | RPA-T4 | anti-human | 50 | Surface | Biolegend | 300554 | B357150 |
| Biotin | IgG4 | N/A | anti-human | 50 | Surface | Invitrogen | A10663 | 2431369 |
| PE-Cy7 | CD8 | SK1 | anti-human | 200 | Surface | BD | 335822 | 1292923+1334364 |

**Secondary staining (sandwich)**

| Fluorochrome | Target | Clone | Reactivity | Dilution | Staining | Company | Catalogus no. | Lot no. |
|---|---|---|---|---|---|---|---|---|
| PE | Streptavidin | N/A | N/A | 100 | Surface | eBioscience | 12-4317-87 | 2699817 |

### Supplementary Table 7 ImageStream flow cytometry panel

**Primary mix**

| Fluorochrome | Target | Clone | Reactivity | Dilution | Staining | Company | Catalogus no. |
|---|---|---|---|---|---|---|---|
| PE-CF594 | CD3 | UCHT1 | anti-human | 50 | Surface | BD | 562310 |
| FITC | PD-1 | MIH4 | anti-human | 25 | Surface | BD | 557860 |
| biotin | IgG4 | N/A | anti-human | 50 | Surface | Invitrogen | A10663 |

**Secondary staining (sandwich)**

| Fluorochrome | Target | Clone | Reactivity | Dilution | Staining | Company | Catalogus no. |
|---|---|---|---|---|---|---|---|
| PE | Streptavidin | N/A | N/A | 25 | Surface | ThermoFisher Scientific | 12-4317-87 |

**Supplementary Table 8 Gene Set Variation Analysis gene sets**

| Gene set | Member genes |
|---|---|
| bMIS_UC (Argmann et al. Gut 2023) | GRIN2D, EBF3, MDFI, MUC5AC, GSDMC, SIX1, SAA2, ICAM1, ANK1, KRT16, CD274, SLC7A5, LY6D, CA9, KRT17, SAA1, MEOX1, GAL, VSIG1, CTHRC1, SERPINB5, ATP5MC1P6, RND1, SMIM25, LINC01270, IL2RA, AC034199.1, NPTX2, HABP2, IGLV6-57, PIM2, IL6, CASP5, CCL11, STC1, SERPINB4, CSNK1A1L, FAP, GABRP, NOTUM, IGLV1-51, IGLV9-49, AC099509.1, TPTE2, IGLV1-47, ALDOB, OSM, TFAP2C, IGLV1-44, IGLV1-40, IGLV1-36, SERPINB3, PDE10A, LINC00473, MRPS31P2, AC017002.3, RPS14P4, AC017002.1, LINC01303, IGKV1-5, IGKV1-9, IGKV3-11, IGKV1-12, IGKV3-15, IGKV3-20, IGLV3-27, TFPI2, GRHL1, DCSTAMP, CASP1P2, IGLV3-25, CEACAM4, LINC02577, ROPN1L, CLDN2, LCN2, IGKV1-39, SERPINB7, LBP, PLEKHS1, DSG3, IGLV3-21, IGKV2D-40, IGHV4OR15-8, IGLV3-19, SLC11A1, RNF183, PADI4, IGLV3-9, MYEOV, IFITM9P, KCNQ3, IGLV2-5, SERPINB2, IGLV3-1, IGLC2, HAS1, C6orf223, AQP9, C4orf50, SOCS1, ALDH1A2, IL1A, AP005233.2, FER1L4, IL1B, CMTM2, NPSR1, CDH3, AL365226.2, FPR1, RPL7P31, NCAPD2P1, FOXQ1, NPSR1-AS1, FPR2, ABCA12, TIMP1, IGF2BP3, FOSL1, SPATA20P1, FIBIN, KCNJ15, EGFL6, UNC5CL, LGALS12, RBMXL2, Z97206.2, Z97206.1, ARNTL2, AC113615.1, AC099524.1, S1PR5, HRH4, AL592164.1, CHRDL2, IL1RN, LINC00114, BGN, TM4SF20, IL13RA2, IGHV1OR15-2, LINC02115, KYNU, AC004585.1, IL11, SPINK4, VNN1, CCNG1P1, LINC01484, MZB1, ESM1, TRIB2, LPL, JSRP1, SYT12, VNN3, CT83, MCEMP1, APOBEC1, S100P, ADGRG3, CCNO, SLC4A11, IGFBP5, DMBT1, AL583785.1, OLFM4, AC112721.2, XKR9, UCN2, C2CD4A, ENPP7, FOLH1, FAM157B, C2CD4B, AC023796.2, ZBP1, LGALS9DP, THBS2, OR52K3P, MLN, CYP24A1, VNN2, IFNE, MIR31HG, MSX2, CCL24, AC115522.1, LYPD5, CHAC1, NOS2, KRT6B, LINC00346, CCR8, NLRP12, HNRNPA1P21, LINC01679, MEFV, CD300E, SIK1, KRT6A, FJX1, COL7A1, TGM2, PCDH17, AL606807.1, INHBB, H19, IL21R, TFF1, HRH2, AFF2, LINC00839, ADM, TRIM29, DCC, CPS1, SMOX, PLAU, SPHK1, GIP, SLCO1B3, SLC6A20, TNFRSF9, LPCAT1, KLK7, KLK6, BCL2A1, PLA2G4E, FCRL5, CD1B, SPNS2, LIPN, LILRA6, ADGRE2, ADGRE3, MNDA, SIRPB1, LINC01993, VWFP1, SOCS3, AC061992.1, IGHV6-1, CCR3, PLEKHN1, RFX8, CXCR2, IGHV1-2, IGHV1-3, TREML2, TREML4, FAM83A, TREM1, IGHV3-11, IGHV1-12, IGHV3-15, IGHV1-18, RPSAP19, LINC01709, DNAJA1P5, OLFM3, CXCR1, KLK10, IGHV3-20, IGHV3-21, IGHV3-23, IGHV1-24, IGHV2-26, IGHV4-28, LINC01819, IGHV4-31, RGS16, LUCAT1, IGHV3-33, IGHV4-34, IGHV4-39, IGHV3-43, IGHV1-45, IGHV1-46, KLK11, WISP1, ITLN2, LILRB2, CPXM1, MAP3K20-AS1, IGHV3-49, FFAR3, DUSP4, CSF2, UBD, OR2I1P, ADAMTS4, TNFAIP6, DEFA6, CXCL8, CXCL6, CXCL1, KLK12, DEFA5, MMP9, INHBA, CXCL5, CXCL3, FFAR2, FAM92B, PTGS2, IGHV5-51, IGHV3-53, IGHV1-58, IGHV4-61, IGHV4-59, STRA6, DUOXA1, CXCL2, FCGR3A, FCGR3B, DUOX2, ABCA13, DUOXA2, CARD6, FCN1, IGHV3-66, IGHV1-69, IGHV2-70D, LILRA5, OLFML2B, ACOD1, ENKUR, TNC, IGHV3-73, SLC7A11, NNMT, AC015969.1, MEDAG, CTLA4, CEMIP, GNA15, APLN, AQP5, PLA2G2A, GCKR, EVA1A, IL17A, PLA1A, CFB, C2, CSF3R, AC007991.4, AC007991.2, IDO1, DKK2, SELP, SELE, FOSB, ZC3H12A, NMUR2, TCN1, CLDN14, AL049836.1, CLEC5A, PI3, ADGRF1, SEMG1, CLDN1, GZMB, MMP7, CD70, COL23A1, MYBPH, CHI3L1, HCAR3, HCAR2, AL354702.1, LAX1, PPP4R4, FCN3, REG1B, PDX1, REG1A, AGT, DEFB4A, CFI, REG3A, ABCG8, ABCG5, ACKR4, PRSS1, PRSS3P1, PROK2, GATA4, PRSS22, FGR, MUC16, NFE2, KCND3-IT1, KCND3-AS1, KCND3, LINC01750, TNFRSF8, LAIR2, FCAR, C4BPB, C4BPA, CD55, CXCL9, GLT1D1, CLDN18, WNT2, AC245128.3, LILRA1, SERPINA3, GPR4, SERPINE1, CHST2, ALDH3B2, TACSTD2, SPP1, IFNG, IL26, IL22, AC084871.2, OLIG1, GPR84, TNIP3, CSF3, ANXA10, GBP1, GBP4, PLA2G3, EPHB1, MMP3, MMP1, MMP10, CXCL10, ART3, GBP5, FCGR1A, CXCL11, SFRP2, AC105046.1, EGR3, LINC02323, COL8A1, ADGRG6, FAM157A, EPHX4, PDZK1IP1, STRIP2, AC021218.1, HTR1D, PDPN, APOL1, AL355483.3, LRP8, AC097478.1, GALNTL6, WNT5A, ALPL, IL21, IL21-AS1, HAPLN3, IGHG4, IGHG2, IGHGP, TDO2, IGHG1, IGHG3, IGHD, ANXA1, REG4, PGLYRP4, S100A9, S100A12, S100A8, S100A3, S100A2, AC244453.2, FCGR1B, TNFRSF10C, KRT80, MIR3945HG, CLEC6A, TESC, CLEC4D, CLEC4E, KRT7, L1TD1, PQLC2L, MYH13, HGF, MYRFL, IL17REL, MLC1, RN7SL471P, RF00019, MIR5571, RN7SL368P |
| Th1 | IFNG, GZMB, IL23, TBX21, IL2, TNF, IL12RB1, IL12RB2, IL18R1, STAT1, STAT4, CXCR3, CCR5, RUNX3 |
| Th2 | IL4, IL5, IL13, GATA3, IL25, IL33, CCL17, CCL22, STAT6, IRF4, BCL6 |
| Th17 | IL17A, IL17F, RORC, CCR6, CCR4, CSF2, STAT3, TNSF11, BATF, RUNX1 |
| Bmem_PC | CD20, CD79A, CD19, EIF4EBP1, TNFRSF13C, IGHM, IGHD, IGKC, IGLC2, CD24, IGHM, VPREB1, PAX5, RSP27 |
| Th17.1_selective (Ramesh et al. JEM 2014) | ME1, ENPP1, ADAM23, COLQ, SLC4A10, CA2, SCRN1, ABCB1, CLEC2B, CCR2, DPP4, KLRB1 |

### Supplementary Figures

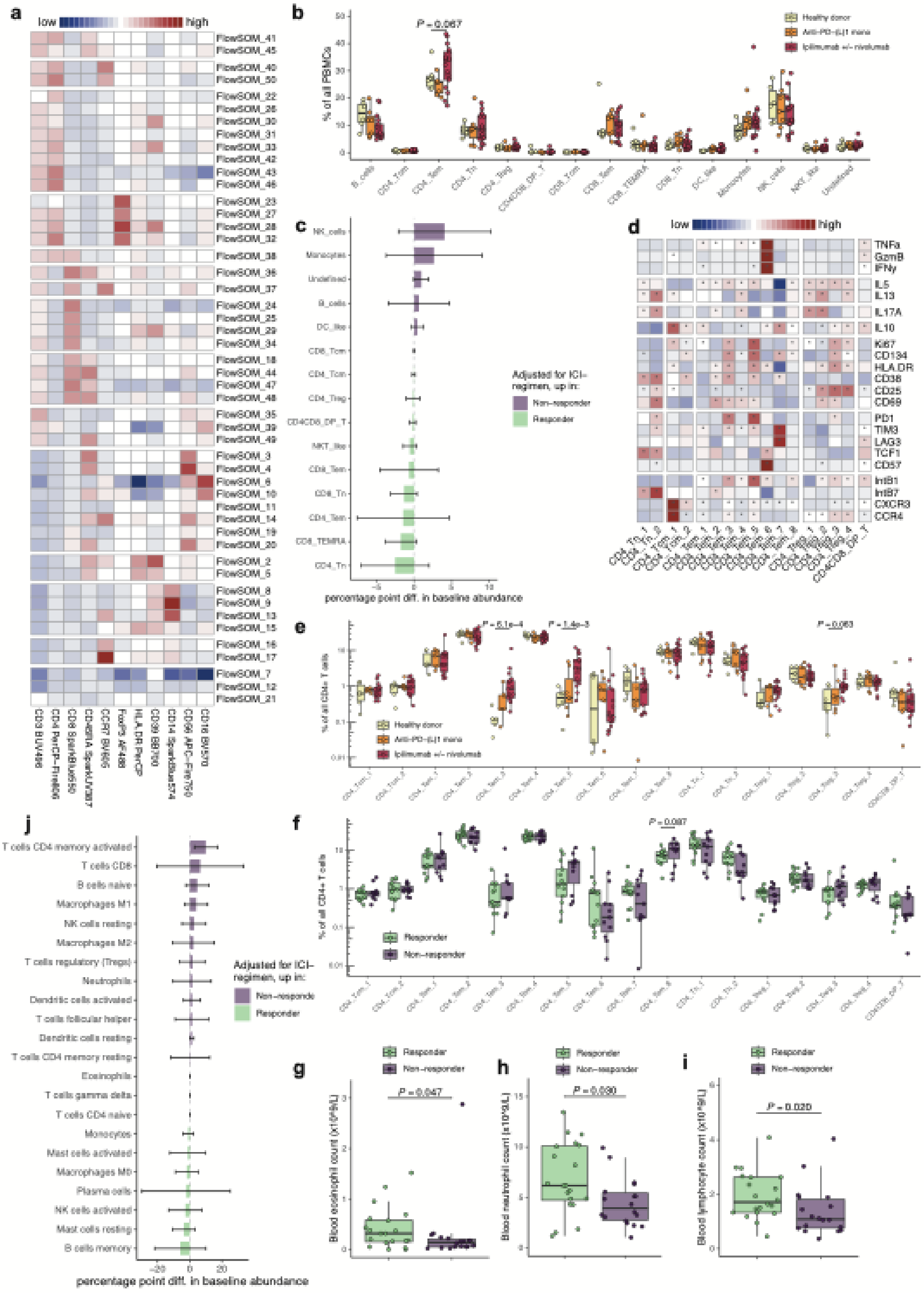

***Supplementary Figure 1 Detailed immunophenotyping of pre-steroids peripheral blood cell subsets.*** *Unadjusted P values are displayed, unless otherwise specified.* ***a*** *Expression heatmap displaying median expression of lineage-defining markers in 49 of 50 FlowSOM metaclusters (FlowSOM metacluster 1 contained debris and was excluded).* ***b*** *Boxplots displaying pre-steroids abundance of different PBMC subsets by ICI treatment type.* ***c*** *Relative pre-steroids abundance of PBMC subsets by response to steroids, adjusted for ICI treatment type. Linear regression coefficients are shown with whiskers indicating 95% confidence intervals.* ***d*** *Expression heatmap displaying median expression of functional markers in $CD4^+$ T cell subsets, grouped from top to bottom as Th1-related, Th2-related, Th17-related, Immunosuppressive, Proliferation/activation, Dysfunction/exhaustion and Homing/trafficking. Except CD57 and HLA-DR, these markers were not use as clustering parameters in FlowSOM. Significant marker enrichment at $P_{adj}<0.05$ is indicated by '*'.* ***e,f*** *Boxplots displaying pre-steroids abundance of $CD4^+$ T cell subsets by (**e**) ICI treatment type or (**f**) response to steroids.* ***g-i*** *Boxplots displaying pre-treatment (**g**) blood eosinophil count, (**h**) blood neutrophil count and (**i**) blood lymphocyte count, by response to steroids in the full extended cohort. Data were missing in 17 patients.* ***j*** *Relative pre-steroids abundance of deconvoluted immune cell types in pre-steroids samples with sufficiently reliable deconvoluted compositions (at $P_{perm}<0.05$), by response to steroids and adjusted for ICI treatment type. Linear regression coefficients are shown with whiskers indicating 95% confidence intervals.*

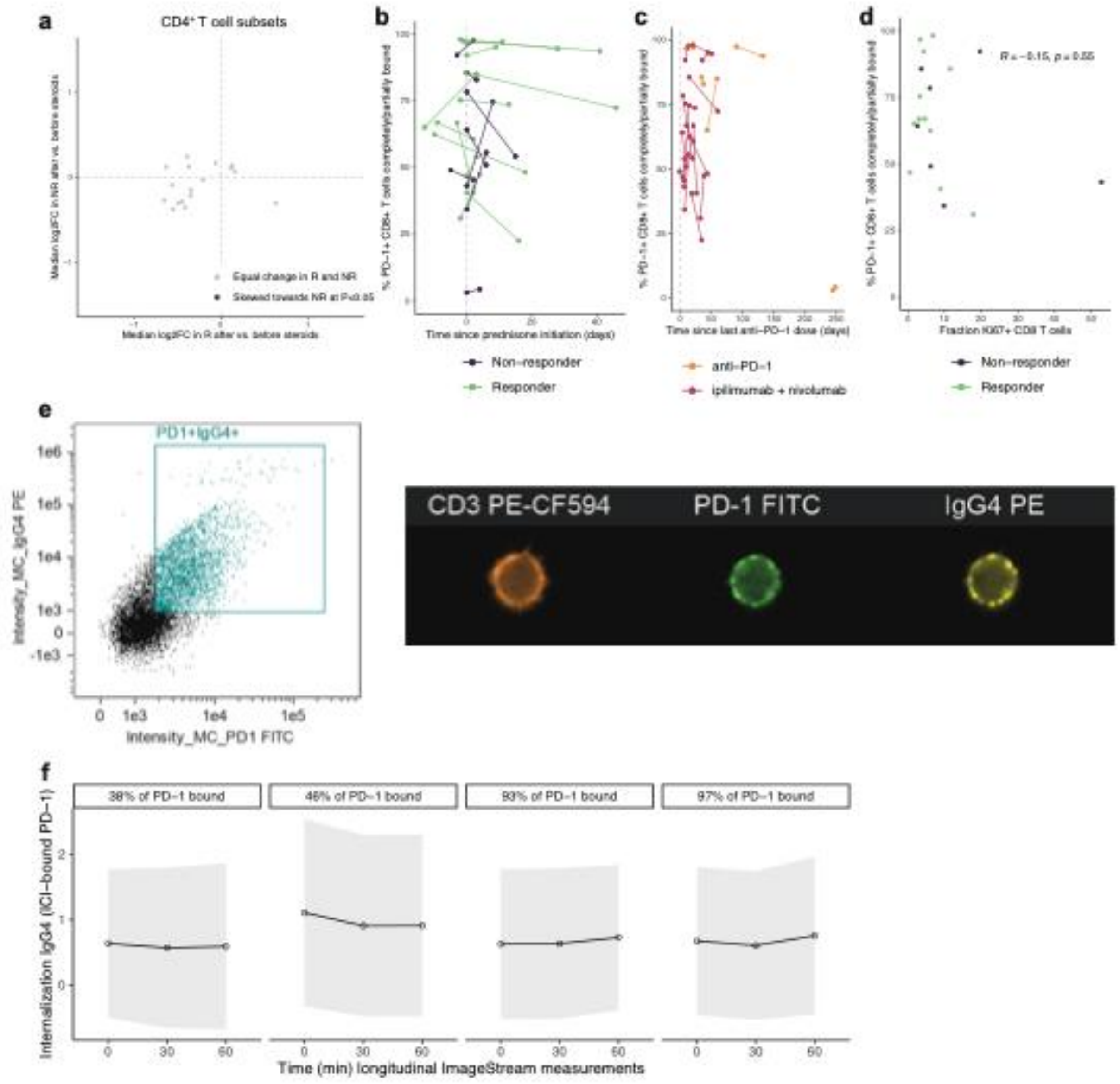


***Supplementary Figure 2 T cell PD-1 receptor occupancy and PD-1 internalization. a*** *Median $log_2$ fold-change of $CD4^+$ T cell subsets after initiation of steroids, in steroid responders (R; x-axis) versus steroid non-responders (NR; y-axis). None of the subsets differed significantly in fold-change between responders and non-responders at unadjusted $P<0.05$. "Skewing towards" an index group can constitute a relative increase in that index group, a relative decrease in the comparator group, or a combination of both.* ***b,c*** *Scatter plots displaying the percentage of PD-$1^+$ $CD8^+$ T cells completely or partially bound by anti-PD-1 therapeutic antibody in relation to (**b**) duration of steroids or (**c**) time since last anti-PD-1 administration, by (**b**) response to steroids or (**c**) ICI treatment type, for paired pre-/on-steroids samples. No formal statistical tests were performed.* ***d*** *Scatter plot displaying the percentage of PD-$1^+$ $CD8^+$ T cells completely or partially bound by anti-PD-1 therapeutic antibody in relation to $CD8^+$ T cell proliferation for pre-steroids samples only. One patient with late-onset toxicity >200 days after the final ICI dose was excluded from this analysis.* ***e*** *Representative images displaying the gate for ICI-bound (PD-$1^+$) T cells and single-cell images showing CD3, PD-1 and IgG4 signal measured with ImageStream.* ***f*** *Internalization of IgG4 (reflecting ICI-bound PD-1) in three longitudinal measurements (0, 30 and 60 min after completion of staining) in four patients, sorted from left to right by the fraction of PD-1 receptor occupancy. Internalization is displayed as the median internalization feature +/- 95% confidence interval, with increasing values over time reflecting net internalization of IgG4 signal.*

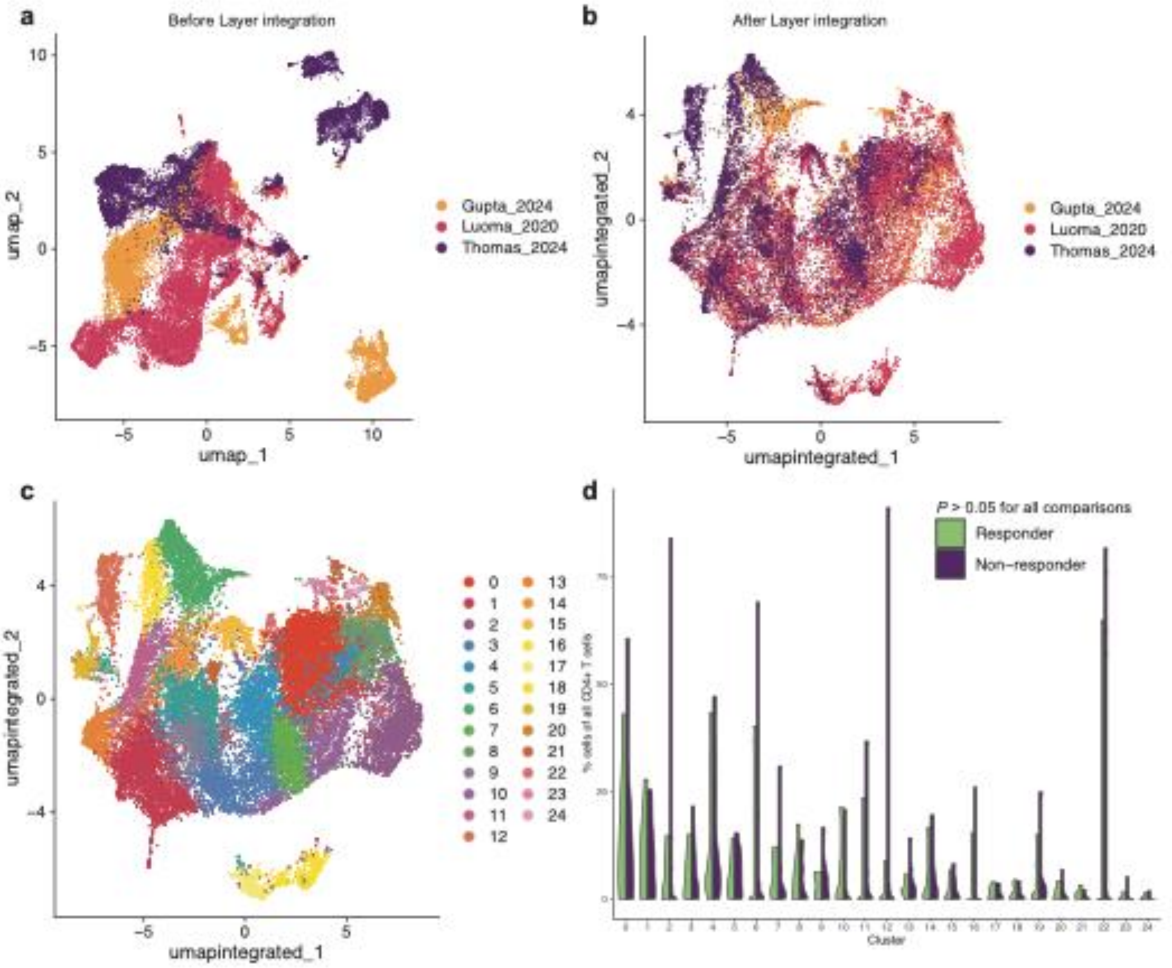


***Supplementary Figure 3 Meta-analysis of CD4⁺ T cell scRNA-seq data. a-c*** *UMAP visualizations of CD4⁺ T cell scRNA-seq data in ICI colitis tissue from three studies (**a**) before and (**b**) after Layer integration to correct for study-specific batch effect and (**c**) after unsupervised clustering of integrated data.* ***d*** *Violin plots displaying the percentage of CD4⁺ T cells in each scRNA-seq cluster, stratified by response to steroids. None of the clusters was differentially abundant between steroid responders and non-responders at unadjusted P<0.05.*

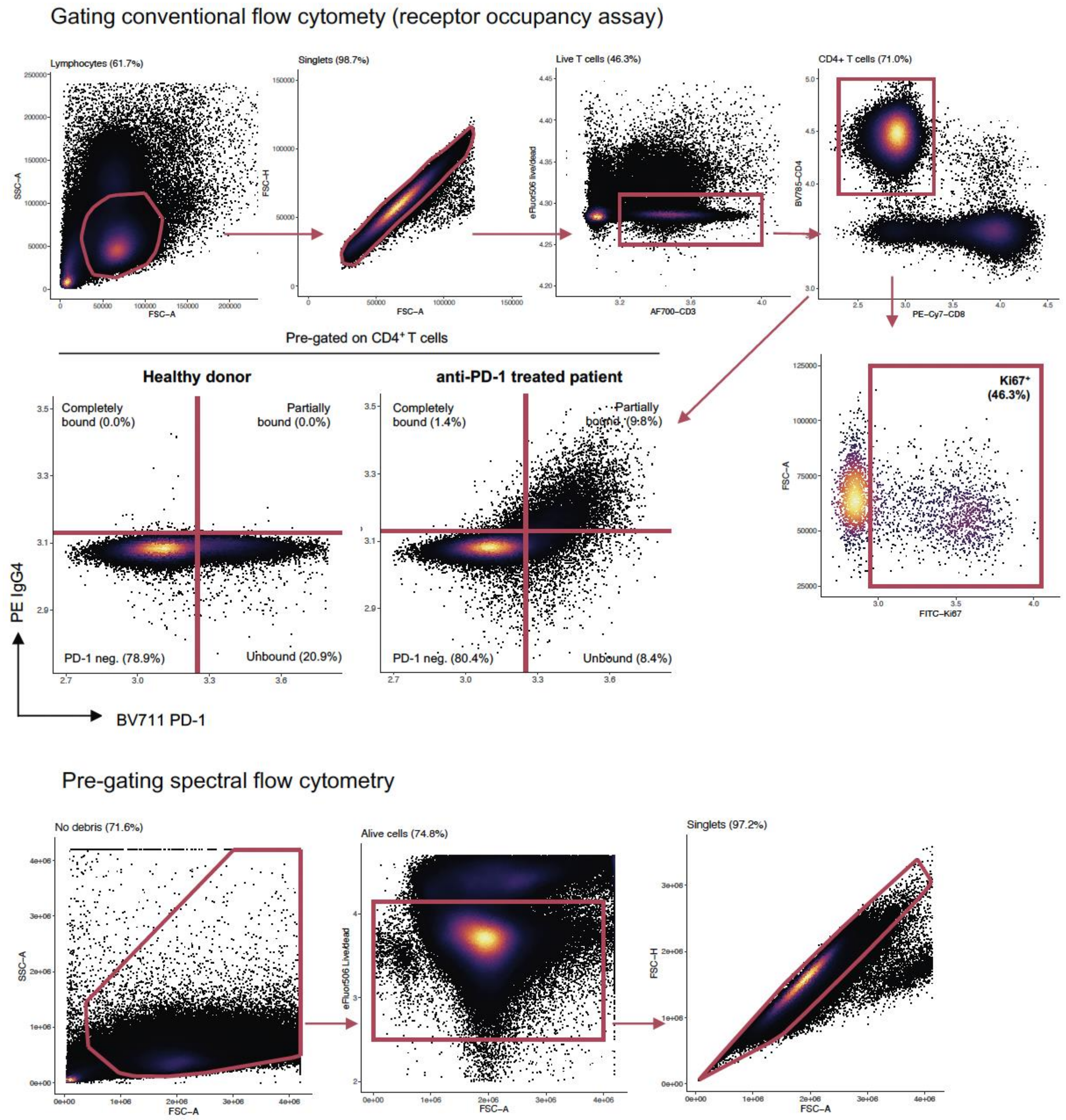


***Supplementary Figure 4 Conventional and spectral flow cytometry gating strategies.*** *Gating strategies in conventional and spectral flow cytometry. For the conventional panel, we consecutively gated for lymphocytes, singlets, live T cells and finally CD4⁺ and CD8⁺ T cells, in which we assessed proliferation and the fraction unbound, partially and completely bound PD-1. For the spectral panel, we pre-gated all non-debris events, live cells and finally singlets. Events were exported as separate .fcs files for further analysis.*
